# A systematic review and meta-analysis of the incidence of *Pneumocystis jirovecii* pneumonia (PJP) in children and young people with cancer, cancer-like conditions or haematopoietic stem cell transplants

**DOI:** 10.64898/2025.12.22.25342677

**Authors:** Connor Evans, Mark Corbett, Chinyereugo Umemneku-Chikere, Helen Fulbright, Bob Phillips

## Abstract

*Pneumocystis jirovecii* Pneumonia (PJP) is a potentially life-threatening fungal infection occurring in immunocompromised individuals, particularly those receiving anti-cancer treatment. However, the incidence of PJP among children and young people (CYP) with cancer, cancer-like conditions or haematopoietic stem cell transplant (HSCT) recipients is unclear. This systematic review aimed to determine the incidence and impact of PJP in this population.

Twelve databases were searched in November 2024. Any study reporting the incidence of PJP in CYP<18 years old with cancer, a cancer-like condition or having received a HSCT were included. Screening, data extraction and quality assessment (using a modified JBI critical appraisal tool for prevalence studies) were conducted in duplicate. Meta-analyses using GLMM methods were used.

Of 7,194 records screened, 106 studies were included. Interpretation of the results of many studies was hindered by limited reporting. For the acute lymphoblastic leukaemia meta-analyses using confirmed events, higher pooled cumulative incidence rates were seen in the no prophylaxis group than in the prophylaxis group (2.86% vs 0.04%). Rates increased to 5.41% in the no prophylaxis group when studies reporting unconfirmed cases were included. Subgroup analyses suggested lower incidence rates in cohorts taking first-line PJP prophylaxis, compared to second-line. The cumulative incidence rate using confirmed events was 0.16% for HSCT patients receiving prophylaxis; data were not available for a no prophylaxis analysis. Only three studies were available for the HSCT total events analysis, producing a rate of 4.27%. Several analyses were subject to some uncertainty due to their high heterogeneity estimates.

PROSPERO registration: CRD42025628682

## Introduction

Children and young people (CYP) with cancer, cancer-like conditions or having received a haematopoietic stem cell transplant (HSCT) are more susceptible to infection. *Pneumocystis jirovecii* pneumonia (PJP) (previously known as *Pneumocystis carinii* pneumonia) is a potentially life-threatening, opportunistic, fungal infection affecting immunocompromised patients, particularly those with low T-cell counts.^1, 2^ Typical symptoms include fever, hypoxia, tachypnoea and severe coughing.^2^

As infections are the most common life-threatening complications in CYP with cancer^3^, preventing infection is an important research priority^4^. Prophylactic antibiotics, such as the combination of trimethoprim (TMP) and sulfamethoxazole (SMX) (TMP/SMX), are usually given to reduce the risk of PJP infection. However, antibiotic use carries the risk of increasing antimicrobial resistance, so good antibiotic stewardship requires an appropriate judgement to be made in balancing these risks.^5^

Though PJP infection in children with cancer is considered rare^6^, estimates of the risk of PJP in CYP undergoing treatment for cancer, cancer-like conditions or receiving HSCTs are uncertain. Therefore, this systematic review was conducted to synthesise the evidence on the rate of PJP infection in these groups.

This systematic review falls under a larger project in collaboration with the Pediatric Oncology Group of Ontario (POGO) to update and develop new clinical practice guidelines for identifying, managing and treating PJP infection in CYP with cancer or children receiving a HSCT. A simultaneous review conducted by the POGO team will synthesise efficacy and safety evidence on treatments for PJP. Together, both reviews will support the development of new guidelines to improve the prevention of PJP in CYP with cancer.

## Methods

This systematic review was conducted according to the protocol registered on PROSPERO (CRD42025628682)^7^ and reported based on PRISMA 2020 guidelines^8^.

### Search strategy

The search strategy was designed by an information specialist (HF) in consultation with the review team. The Ovid MEDLINE strategy included subject headings and free-text terms and was adapted for the other databases. Two concepts were included in the strategy: *pneumocystis jirovecii* pneumonia and children and young people. No study design, date or language restrictions were applied.

On 21^st^ November 2024 the following databases were searched: MEDLINE®; EMBASE; PsycINFO; Maternity & Infant Care Database; KSR Evidence; ECRI Guidelines Trust; Cochrane Database of Systematic Reviews; Social Sciences Citation Expanded; ProQuest Dissertations & Theses Citation Index; CINAHL Ultimate; PROSPERO; and the International HTA Database. The full search strategies are reported in Supporting Methods S1.

Reference lists of included studies and relevant systematic reviews were also checked. Non-English language studies were screened using a translation application on a mobile device.

### Eligibility criteria and screening

Studies were independently screened by two researchers (CE, MC) with disagreements resolved via team discussion using the following eligibility criteria:

#### Population

CYP<18 years old with cancer, a cancer-like condition or having received any HSCT. Studies reporting results for children and adults together were only included when at least 90% of the population were CYP<18 years.

#### Exposure

Suspected or confirmed diagnosis of PJP infection.

#### Outcomes

Studies had to include the rate of PJP infection including prevalence and/or incidence to be eligible. Other outcomes were extracted only when a study reported PJP rate, but included: timing of infection, PJP severity, PJP mortality, and PJP risk factors.

#### Study design

Prospective or retrospective cohort or cross-sectional studies were eligible. Case-control studies and case reports were excluded, as were studies where the whole population were autopsies and the study only reported incidence of PJP post-death.

### Data extraction

Data were extracted by one researcher (CE) and independently checked by another (MC, CC) with disagreements resolved via team discussion. PJP diagnoses based on bronchoalveolar lavage, lung biopsy specimens, computerised tomography or chest X-ray changes in a clinical situation with recovery as expected, were considered as confirmed cases. Diagnoses based on reverse-transcription polymerase chain reaction blood tests alone were considered unconfirmed, as were cases described as ‘suspected’ and cases where there was no diagnosis description. Graphical data were estimated from digitised plots using the PlotDigitizer software online (https://plotdigitizer.com/app). Non-English language studies were extracted by a researcher fluent in that language.

Many more studies were included in the review than was anticipated, so a pragmatic approach was taken to data extraction and quality assessment. Studies with fewer than 100 HSCT or acute lymphoblastic leukaemia (ALL) prophylaxis patients were extracted using a basic data extraction template without quality assessments, and their data were not included in the meta-analyses. Studies with mixed cancer populations that did not report PJP incidence by cancer type were also of limited value and received basic data extraction. These studies were not included in the meta-analyses because of the expected scarcity of events or low clinical interpretability of heterogenous risks.

### Quality assessment

Quality assessments were conducted by one researcher (CE) and independently checked by another (MC, CC) using a modified version of the Joanna Briggs Institute critical appraisal tool for prevalence studies.^9^ Disagreements were resolved via team discussions. The modified tool is presented in Supporting Methods S2.

### Data synthesis

The primary meta-analyses were performed using random effects generalized linear mixed models (GLMM) via the metaprop package in RStudio using the logit-link function^10^; fixed effect analyses were also performed. Secondary meta-analyses were also undertaken using the fixed effect beta-binomial (BB) model via the oad package in RStudio, and Bayesian random effects BB model via WinBUGS through the Rjags package in RStudio (see Supporting Methods S3 for further details).^10^ These meta-analysis methods avoid the use of a continuity correction when there are no events, which are likely to produce biased estimates.^11^ Although the forest plots display 95% confidence intervals (CIs) for studies with no events, these were calculated only for ease of visualisation, using the continuity correction method.^12, 13^ They were not used in the actual meta-analyses (which instead used the GLMM structure).

Analyses were conducted and presented according to whether PJP events were classed as: i) confirmed events, based on the diagnostic methods reported (a ‘confirmed event’ analysis) or ii) total events, any reported PJP event (including suspected events) regardless of the level of detail reported on diagnostic methods (a ‘total event’ analysis). Studies with no PJP events were included in the confirmed event meta-analyses. Analyses were also conducted based on the line of PJP prophylaxis, where this was reported (or could be reasonably assumed) using total events, to maximise the limited available evidence. Studies which could not be included in a meta-analysis were tabulated and summarised narratively.

## Results

### Study selection

The database searches identified 7,194 records (after deduplication), with 222 full-texts screened. Of these, 109 met the inclusion criteria and were included in the review (see Supplemental Table S1 for studies excluded during full-text screening). Five further studies were identified from searching reference lists of included studies and relevant reviews. In total, 106 studies (114 records) were included in the review (see Figure 1): 66 were extracted in full (Supplemental Table S2) and 43 studies had basic data extraction (Supporting Results S3); three studies had both.

**FIGURE 1.**
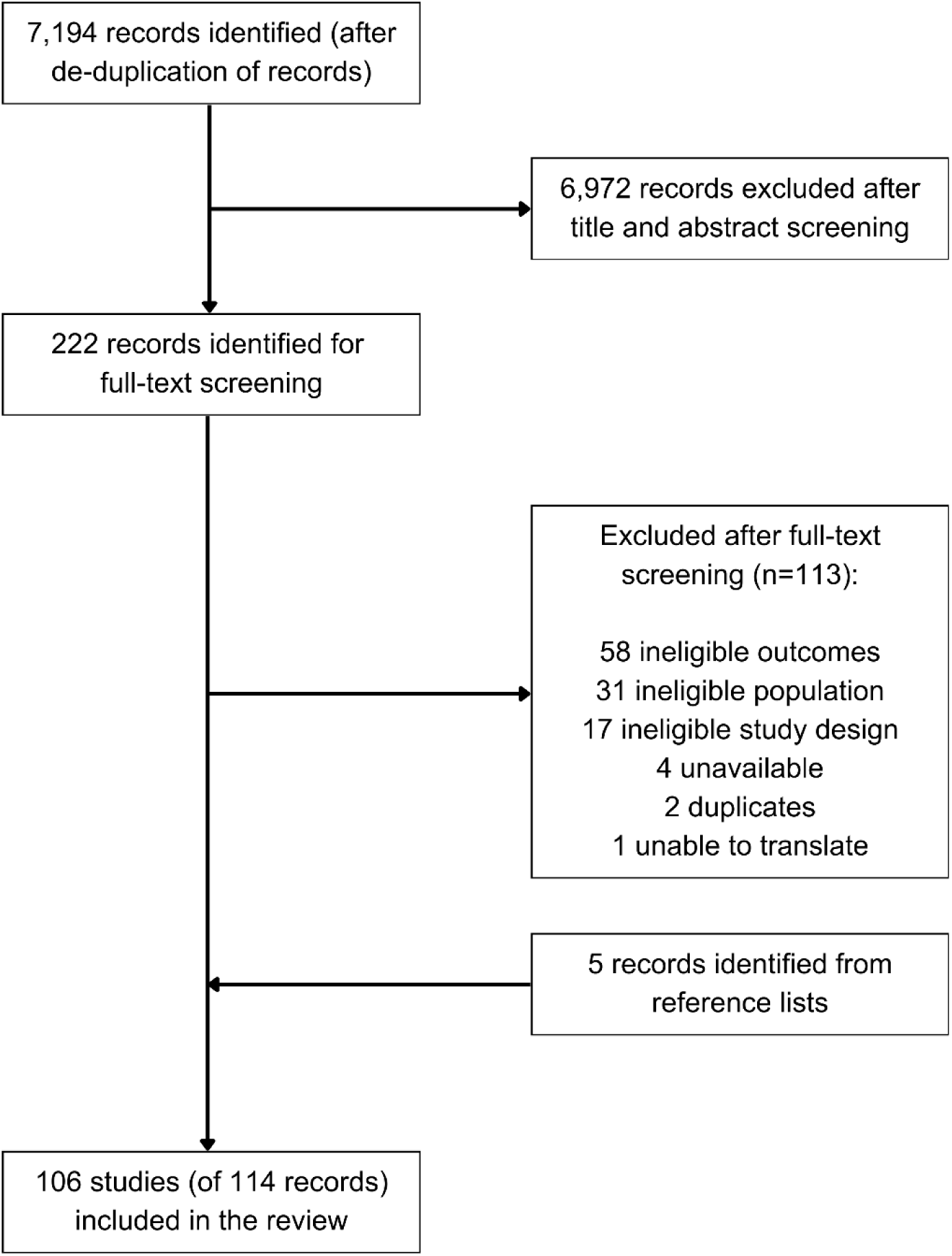
PRISMA flow diagram.

### Quality assessment

Sixty-six studies were quality assessed^14–79^ (Table 1). Studies were limited by their reporting, although in 23% (15/66) of studies, assessment of PJP incidence was not either a stated or implied aim of the study. Although most studies (54/66, 82%) appeared to recruit an appropriate target population, over half (35/66, 53%) did not describe participant characteristics and settings in adequate detail. Only 42% (28/66) of studies clearly reported using valid methods for the detection (and diagnosis) of PJP.

**TABLE 1.**
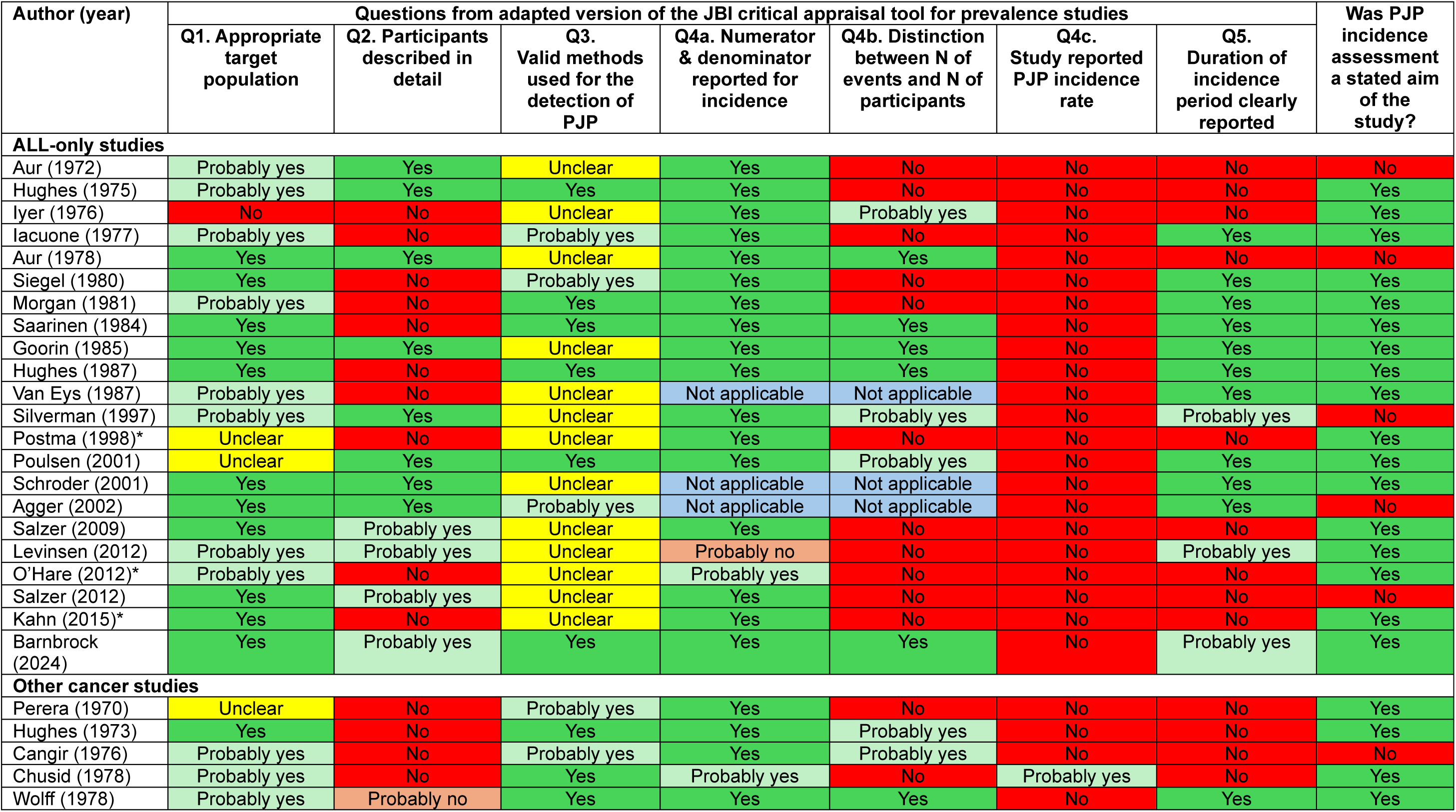
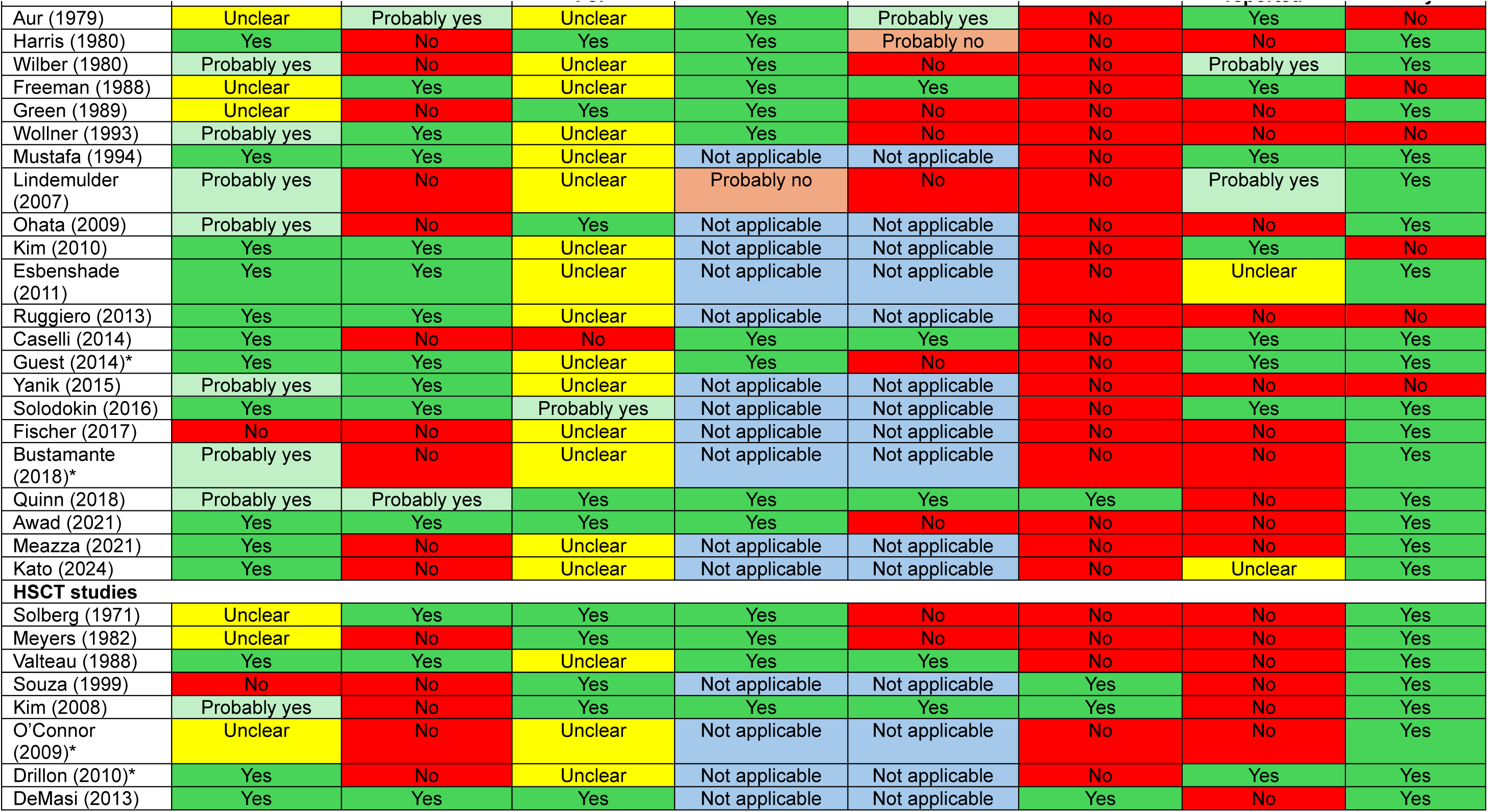
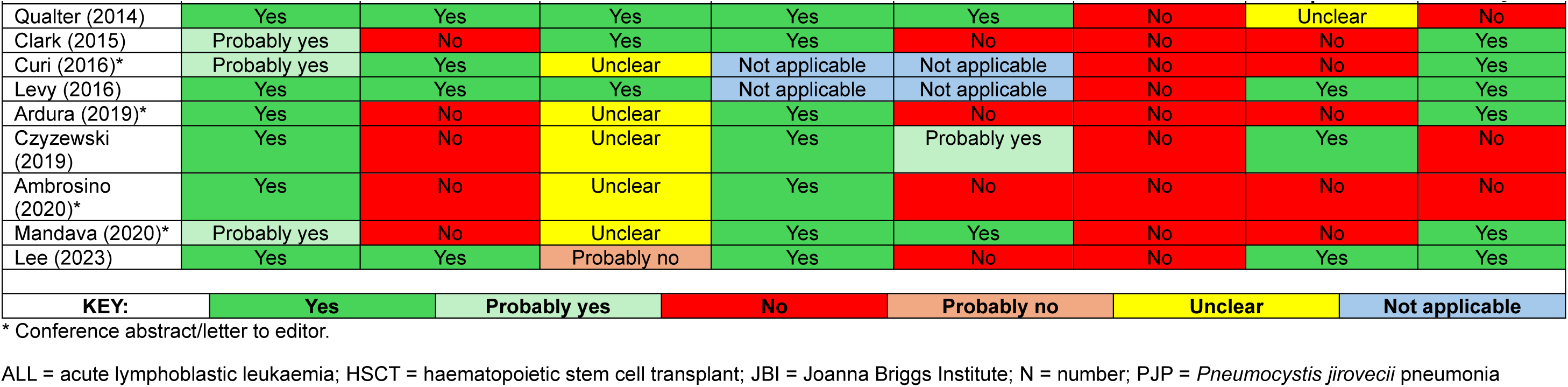
Quality assessment results.

In the 46 studies which reported at least one case of PJP, all but two of them clearly reported the numerator and denominator to allow calculation of incidence proportions. Just 43% (20/46) of those (PJP event) studies reported data to allow a distinction between the number of events (either of PJP or of HSCT) and the number of individuals, which is necessary to avoid double-counting of individuals in either numerators (PJP cases) or denominators (HSCT patients). Only five studies reported, or stated an aim to report (if there were no cases), a PJP incidence rate (i.e. an analysis which takes into account losses to follow-up and when PJP occurs), as well as (or instead of) a simple incidence proportion.^36, 56, 63, 64, 67^ An indication of the duration of the period when PJP cases were recorded was reported in only 41% (27/66) of studies.

### Synthesis

#### Study and participant characteristics

Study and participant characteristics for studies with full data extraction are presented in Supplemental Table S2. Most studies were of mixed cancers or ALL-only cohorts.

##### HSCT studies

Eighteen HSCT studies had full data extraction^56, 60–76^, dating from 1971-2023. Where reported, most were retrospective cohort studies (17 studies, 94%)^56, 61–76^ conducted in the USA (15 studies, 83%)^56, 60, 61, 63–72, 74, 75^, which recruited from one treatment centre (14 studies, 78%)^56, 60, 62–71, 74, 76^. Where reported, a total of 14,497 patients receiving at least one HSCT were included. Sample sizes ranged from nine to 11,056, with half the studies including 100-200 CYP. The mean or median age ranged from 4.5 to 11.7 years (though only reported in eight studies^60, 62, 67–71, 76^). Specific information on age and sex was generally limited. Most studies (12, 67%) were of allogeneic HSCTs.^60, 61, 63, 64, 66–68, 70, 71, 73, 75, 76^ Most HSCTs were given for malignant diseases (particularly leukaemia). Fifteen studies included a cohort receiving PJP prophylaxis^56, 63–76^, with pentamidine being the most common type of first- or second-line prophylaxis (where reported).

##### ALL-only studies

Twenty-two studies assessing only CYP with ALL had full data extraction^14–32, 77–79^, dating from 1972-2024. Where reported, most studies were retrospective cohort studies (12 studies, 55%)^16, 17, 20, 21, 25, 27–31, 77, 78^, conducted in the USA (14 studies, 63%)^14–20, 22–25, 30, 31, 79^, which recruited from one treatment centre (11 studies, 69%)^14–18, 20, 21, 23, 26, 27, 78^. A total of 10,610 patients were included across the 22 studies. Where reported, median age ranged from seven months to 5.2 years (across seven studies^15, 18, 22, 25, 27, 28, 32^). Three studies only recruited infants under one years old.^25, 30, 79^ Across the ten studies reporting sex, 57% were male.^14, 15, 18, 21, 22, 25, 27, 29, 32, 77^ Twelve studies included at least one cohort receiving PJP prophylaxis^20, 23, 24, 26, 28–32, 77–79^, with TMP/SMX being the most common type.

##### Other cancer studies

Twenty-seven studies reported on mixed cancer populations (i.e. with varied diagnoses)^33–59^, dating from 1970-2024. Where reported, most studies were retrospective cohort studies (19 studies, 70%)^33, 34, 36–40, 43, 45, 46, 48, 51, 53–59^ conducted in USA (21 studies, 78%)^33–45, 48, 51–57^ which recruited from one treatment centre (17 studies, 77%)^33, 34, 36–38, 40, 43–46, 48, 53–57, 59^. A total of 45,435 CYP were included: 40% (n=18,113) with haematological cancers/lymphoma, 38% (n=17,280) with sarcomas/solid tumours, 20% (n=9,163) with brain(stem)/CNS tumours, and 2% (n=879) with other cancers. Where reported, median age ranged from 5.5 years to 12 years (across nine studies^38, 41, 43, 44, 47, 49, 52, 53, 56^). Across the ten studies reporting sex^38, 41, 43, 44, 47–49, 51, 52, 56^, 54% were male. Sixteen studies included at least one cohort receiving PJP prophylaxis.^39, 44–53, 55–59^ TMP/SMX was the most common type of prophylaxis received, followed by pentamidine.

#### Incidence of PJP - Meta-analysis results

##### PJP incidence in children who have received a HSCT

For the HSCT population which received a prophylactic treatment to prevent PJP, the pooled cumulative incidence rate for the confirmed event analysis was 0.17% (95% CI: 0.03 to 0.95, 10 studies, Table 2); Figure 2 shows there were only four PJP events in total (over three studies), with the other seven studies all reporting no PJP events. The heterogeneity estimate was low (I^2^=0%). For the total events analysis, the rate increased to 0.48% (95% CI: 0.22 to 1.02, 15 studies, Supporting Results S1, Figure 2). Although only six of the 15 studies in this analysis had no PJP events, the I^2^ heterogeneity estimate remained at 0%, despite the forest plot appearing to be visually more heterogeneous (than the confirmed events analysis). The total events analysis was dominated by one very large study by Ardura et al^72^ which had 130 PJP events in 11,056 children. Despite its large sample size, it was unclear which methods were used to identify events in this study so its PJP incidence (1.18%) is subject to some uncertainty.

**FIGURE 2.**
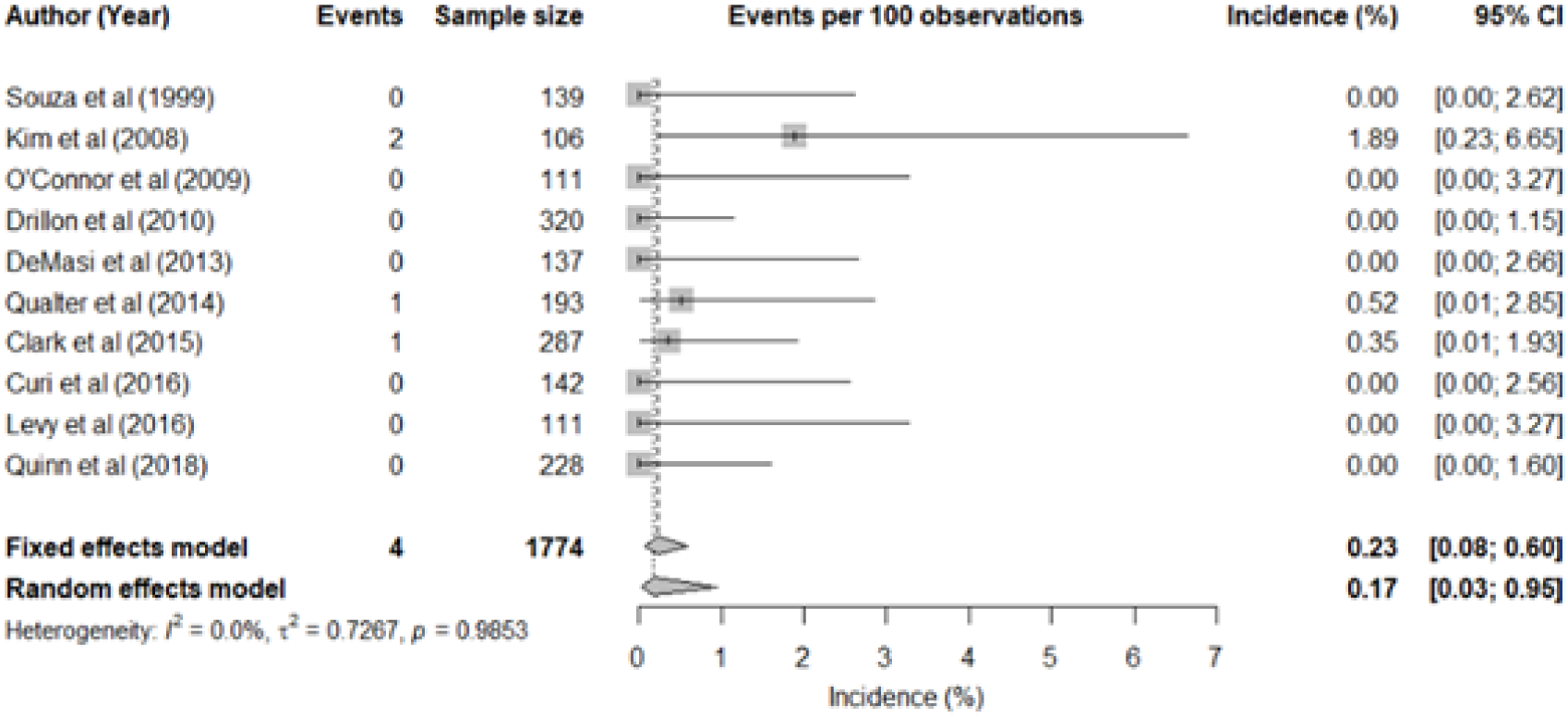
Forest plot presenting incidence rates of confirmed PJP events in HSCT cohorts receiving prophylaxis.

**TABLE 2.** Meta-analysis results of PJP incidence proportion data (using GLMM random effects analyses)

| Population and Prophylaxis Group | PJP cumulative incidence % |  |  |  |
| --- | --- | --- | --- | --- |
|  | No. of studies, I<sup>2</sup> | Using confirmed events (95% CI) | Using total events (95% CI) | No. of studies, I<sup>2</sup> |
| Prophylaxis cohorts |  |  |  |  |
| HSCT | 10, 0% | 0.17 (0.03 to 0.95) | 0.48 (0.22 to 1.02) | 15, 0% |
| ALL | 14, 17% | 0.04 (0.003 to 0.46) | 0.49 (0.19 to 1.26) | 20, 83% |
| AML | NA | NA | 1.16 (0.75 to 1.79) | 3, 0% |
| No prophylaxis cohorts |  |  |  |  |
| HSCT | NA | NA | 4.27 (0.85 to 18.75) | 3, 78% |
| ALL | 12, 68% | 2.86 (1.46 to 5.51) | 5.41 (3.12 to 9.21) | 19, 85% |
| Neuroblastoma | 4, 0% | 3.33 (1.60 to 6.83) | - | - |
| Rhabdomyosarcoma | NA | NA | 1.74 (0.47 to 6.24) | 3, 0% |
| Wilm's Tumour | NA | NA | 0.76 (0.22 to 2.62) | 3, 1% |

NA - Data not available for a meta-analysis
ALL = acute lymphoblastic leukaemia; AML = acute myeloid leukaemia; CI = confidence interval; GLMM = generalized linear mixed model; HSCT = haematopoietic stem cell transplant; NA = not applicable; PJP = *pneumocystis jirovecii* pneumonia

For children receiving first-line PJP prophylaxis the pooled cumulative incidence rate (total events analysis) was 0.15% (95% CI: 0.04 to 0.60, 5 studies, Supporting Results S1, Figure 4). The second-line prophylaxis rate was higher, being 0.54% (95% CI: 0.20 to 1.42, 5 studies, Supporting Results S1, Figure 5). Data from cohorts who had not received prophylaxis were only reported in three studies. In a total events analysis, the pooled rate was 4.27% (95% CI: 0.85 to 18.75, Supporting Results S1, Figure 1) though individual estimates varied widely, which was reflected in the heterogeneity estimate (I^2^=78%). Data were not available for a confirmed events analysis.

##### PJP incidence in children with ALL

For the ALL population which received a prophylactic treatment to prevent PJP, the pooled cumulative incidence rate for the confirmed event analysis was 0.04% (95% CI: 0.003 to 0.46, 14 studies, I^2^=17%, Table 2); Figure 3 shows there were five PJP events in total (across two studies), with the other 12 studies all reporting no PJP events. For the prophylactic treatment total events analysis, the rate increased to 0.49% (95% CI: 0.19 to 1.26, 20 studies, I^2^=83% Supporting Results S2, Figure 5).

**FIGURE 3.**
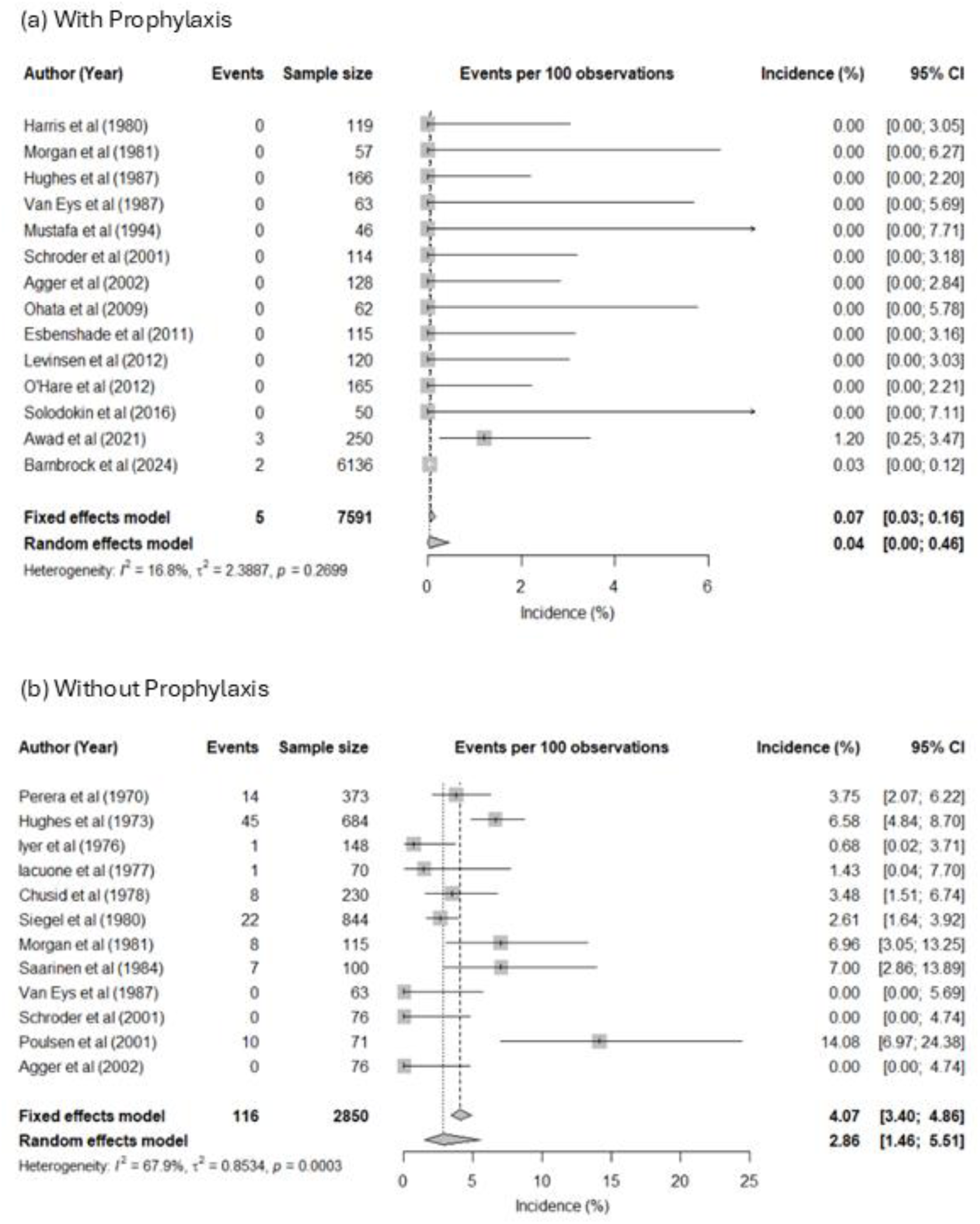
Forest plots presenting incidence rates of confirmed PJP events in ALL cohorts receiving a) prophylaxis, b) no prophylaxis.

For children receiving first-line PJP prophylaxis the pooled cumulative incidence rate (total events analysis) was 0.29% (95% CI: 0.13 to 0.66, 13 studies, I^2^=0%, Supporting Results S2, Figure 7). The second-line PJP prophylaxis rate was 0.43% (95% CI: 0.06 to 2.97, 3 studies, I^2^=0%, Supporting Results S2, Figure 8); though this analysis is limited since it utilises data from only three small cohorts; there was also only one PJP event in total, so caution is needed when interpreting this result.

The analysis of children who had not received prophylaxis produced a pooled cumulative incidence rate using confirmed events of 2.86% (95% CI: 1.46 to 5.51, 12 studies); the heterogeneity estimate was high (I^2^=68%, Figure 3). This appears to be driven by the high rate of PJP (14%) in Poulsen et al^27^, during a steroid-containing maintenance phase. In the no prophylaxis total events analysis, the rate increased to 5.41% (95% CI: 3.12 to 9.21, 19 studies, I^2^=85%, Supporting Results S2, Figure 1) and heterogeneity also increased.

Results for the sensitivity analyses were very similar to the main analyses (Supporting Results S2, Table 1). Results for the secondary meta-analyses are reported in Supporting Results S1 and S2.

##### PJP incidence in children with other types of cancer

Data from four other populations were suitable for meta-analysis. In children with acute myeloid leukaemia receiving PJP prophylaxis a total events analysis produced a rate of 1.16% (95% CI: 0.75 to 1.79, 3 studies, I^2^=0%, Supporting Results S2, Table 2 and Figure 9) with the analysis being dominated by one large study.^51^ The other three meta-analyses were all for no prophylaxis cohorts, resulting in rates of 3.33% (95% CI: 1.60 to 6.83, 4 studies, I^2^=0%) in children with neuroblastoma using confirmed events, 1.74% (95% CI: 0.47 to 6.24, 3 studies, I^2^=0%) in children with rhabdomyosarcoma using total events, and 0.76% (95% CI: 0.22 to 2.62, 3 studies, I^2^=1%) in children with Wilm’s tumour using total events (Supporting Results S2, Table 3, Figures 10-12).

**TABLE 3.** Total number of PJP events per disease type.

| Cancer type | Number of studies | Total sample size | Total number of PJP events<sup>a</sup> |
| --- | --- | --- | --- |
| **Haematological cancers/lymphoma** |  |  |  |
| Acute lymphoblastic leukaemia | 31 | 19,754 | 348 |
| Acute myeloid leukaemia | 6 | 1,832 | 20 |
| Non-Hodgkin's lymphoma | 6 | 2,423 | 18 |
| Hodgkin's lymphoma | 5 | 1,540 | 6 |
| Lymphoma, NOS | 5 | 381 | 11 |
| Leukaemia/lymphoma | 3 | 1,586 | 3 |
| Leukaemia, NOS | 3 | 1,091 | 47 |
| Haematological cancers, NOS | 2 | 109 | 0 |
| Aplastic anaemia | 1 | 7 | 0 |
| **Sarcoma/solid tumours** |  |  |  |
| Solid tumours, NOS | 10 | 13,473 | 30 |
| Neuroblastoma | 5 | 706 | 7 |
| Rhabdomyosarcoma | 3 | 588 | 6 |
| Wilms tumours | 3 | 846 | 4 |
| Ewing's sarcoma | 2 | 469 | 1 |
| Sarcoma, NOS | 2 | 273 | 0 |
| Retinoblastoma | 1 | 274 | 0 |
| Osteosarcoma | 1 | 331 | 0 |
| Germ cell tumour | 1 | 188 | 0 |
| Hepatoblastoma | 1 | 101 | 0 |
| Nasopharyngeal carcinoma | 1 | 31 | 0 |
| **Brain tumours** |  |  |  |
| Brain/CNS tumours, NOS | 5 | 9,106 | 15 |
| Brainstem tumours/ DIPG | 3 | 57 | 1 |
| **Other** |  |  |  |
| HSCT recipients | 18 | 14,608<sup>b</sup> | 167 |
| 'Other'/'Other malignancies' | 4 | 45 | 0 |
| Langerhans histiocytosis | 4 | 319 | 3 |
| Rare tumours, NOS | 1 | 167 | 0 |
| Solid tumour/neuro-oncology, NOS | 1 | 348 | 2 |

<sup>a</sup> Total number of PJP events is the number of confirmed, suspected and unclear events combined and is irrespective of prophylaxis use.
<sup>b</sup> Includes one study that reported the number of HSCTs, rather than the number of patients receiving HSCT (n=111 HSCTs).
CNS = central nervous system; DIPG = diffuse intrinsic pontine glioma; HSCT = haematopoietic stem cell transplant; NOS = not otherwise specified; PJP = *pneumocystis jirovecii* pneumonia

#### Incidence of PJP – other cancer studies not included in a meta-analysis

Results for all studies with full data extraction included in the review are reported in Supplemental Tables S3-5. Five prophylaxis studies of children with brain or brainstem tumours all reported no PJP events^46, 47, 49, 53, 58^, there was also one study where no prophylaxis was given - Freeman et al^41^ reported one event in 34 patients (3%). Cohorts for other cancers were typically small (Supplemental Table S5), the exception being the large prophylaxis study by Guest et al which reported total event rates of 0.3% in children with Hodgkin lymphoma, 0.4% in non-Hodgkin lymphoma and 0.2% in CNS cancers.^51^

Table 3 presents the total number of studies, patients and PJP events by diagnosis across all 66 studies with full data extraction, to highlight the differing volumes of available evidence across diseases. Though numbers of PJP events were relatively small across nearly all cancer diagnoses, there was a higher proportion of events among patients with haematological cancers compared to patients with solid tumours or brain tumours.

Results for other outcomes are reported in Supporting Results S4 and Supplemental Tables S3-5. Results for studies with basic data extraction are presented and summarised in Supporting Results S3.

## Discussion

### Summary of review findings

Our review included 106 studies reporting data on the rate of PJP infection in CYP with cancer, cancer-like conditions or receiving HSCT. Although the review eligibility criteria were broad, the reported data related largely to ALL and HSCT cohorts. Most of the data used in our meta-analyses were derived from retrospective single-centre studies, conducted in the USA. In children receiving a prophylactic treatment to prevent PJP, pooled cumulative incidence rates for our confirmed event meta-analyses were very low, ranging from 0.04% (ALL) to 0.16% (HSCT). Total event analysis rates increased to around 0.5% for both ALL and HSCT. Subgroup analyses, based on line of prophylaxis suggested lower incidence rates in cohorts taking first-line PJP prophylaxis (when compared to second-line). However, the reliability of these results is limited by the small number of available studies and the use of total events.

Many of the cohorts not receiving prophylaxis were studied in the 1970s and 1980s. For the confirmed case ALL analyses, higher rates were seen in the no prophylaxis group than in the prophylaxis group (2.86% vs 0.04%). Rates increased to 5.41% in the total events analysis. For HSCT, only three studies were available for the total events analysis (with a 4.27% rate); no data were available for a no prophylaxis confirmed case meta-analysis.

The results of several analyses are subject to some uncertainty due to their high heterogeneity estimates. Greater heterogeneity was notable in our total events analyses since they also included studies where confirmation methods were either not optimal or unclear. Heterogeneity in our no prophylaxis analyses (in particular) may also be driven by study variation in follow-up durations and in phases of treatment (e.g. induction versus maintenance phase). We also acknowledge that our heterogeneity estimates may be more difficult to interpret since several meta-analyses include many studies with no events; it is unclear how meaningful I^2^ estimates are in these scenarios. There was no evidence to suggest changes in PJP prevalence over time (in either the prophylaxis or no prophylaxis groups). Nonetheless, our most reliable analyses (which used confirmed events) were consistent in showing very low rates of PJP across both ALL and HSCT where prophylaxis was used.

It is also important to note that PJP cases were often reported as having occurred in patients who had discontinued their prophylactic treatment.^26^ ^32^ ^45^ For example, Postma et al reported that in all 14 patients with PJP, prophylaxis had been discontinued 1 to 16 months previously (median four months). However, many of the remaining studies which reported numerous PJP events in prophylaxis cohorts were only reported as conference abstracts^31^ ^51^ ^72^ or as a letter to the editor,^26^ which limited further investigation into this observation.

### Implications for future research

Our quality assessment results highlighted several issues which should be considered in future studies. Reporting issues were common, particularly with respect to: the diagnostic methods used to confirm PJP cases, the duration of the period when PJP cases were recorded, and distinguishing between individuals and events (i.e. avoiding double-counting patients). Our meta-analysis results are limited in that they are based on cumulative incidence (incidence proportion) data. Future studies are needed to provide more accurate results by reporting PJP incidence rates, rather than proportions, to account for both losses to follow-up and when PJP occurs (and proportions do not).

### Strengths and limitations

Our systematic review was undertaken using transparent and robust methods. The bibliographic database searches were comprehensive, and our inclusion criteria were broad, which enabled identification of all relevant studies. Thorough quality assessments (with review-specific questions) were made to evaluate limitations across the evidence-base. The methods used in our meta-analyses ensured that results from the abundance of studies without PJP events did not skew our pooled estimates.

The main limitation of our review was the evidence identified. We included studies which did not report a stated aim of estimating PJP incidence since they might produce different results than studies which did. Many studies were limited by lack of reporting on prophylaxis use; rather than excluding these data, we used clinical judgement (BP) to classify studies as being prophylaxis or no prophylaxis cohorts, based on both study year and clinical characteristics.

### Conclusions and implications for practice

This review summarises the rates of PJP infection associated with therapies for cancer and conditioning for HSCT, with observational data confirming the effectiveness of delivering prophylaxis. Knowing these rates of infection will allow clinicians, families and patients to make more informed, shared decisions about ongoing antibiotic treatment.

## Supporting information

Supporting Results S1

Supporting Results S2

Supporting Results S3

Supporting Results S4

## Competing interests

The authors have no competing interests to report.

## Acknowledgements

We acknowledge the help of David Marshall and Jennifer Brown in translating non-English language papers included in the review. We also thank Mark Simmonds for providing advice with the statistical analyses.

This systematic review was funded by CCLG/Candlelighters (Grant number: CANSC20227).

## Data availability statement

All data included in the systematic review is already available in the public domain.

## Notes

### Competing Interest Statement

The authors have declared no competing interest.

