## Supplemental Table S1 for "A systematic review and meta-analysis of the incidence of *Pneumocystis jirovecii* pneumonia (PJP) in children and young people with cancer, cancer-like conditions or haematopoietic stem cell transplants"

**Supplemental Table S1. List of studies excluded at full-text screening and reasons for exclusion**

| **Author (Year)** | **Title** | **Reason for exclusion** |
| --- | --- | --- |
| Al-Faris (2007) | Does consolidation with autologous stem cell transplantation improve the outcome of children with metastatic or relapsed ewing sarcoma? | Wrong outcome |
| Araki (1992) | Japanese clinical statistical data of patients with Pneumocystis carinii pneumonia | Wrong study design |
| Azeez (2020) | Sequencing and phylogenetic of dihydropteroate synthase (Dhps) gene in Pneumocystis jirovecii isolated from immunocompromised patients in Iraq | Wrong population |
| Bachmann (1953) | The presence of Pneumocystis carinii in interstitial plasma cell pneumonia in infants | Wrong study design |
| Barrett (1983) | Interstitial pneumonitis following bone marrow transplantation after low dose rate total body irradiation | Wrong population |
| Bashey (1990) | Pneumocystis prophylaxis after bone marrow transplantation for severe aplastic anaemia | Wrong study design |
| Beneduce (2022) | Blinatumomab in Children and Adolescents with Relapsed/Refractory B Cell Precursor Acute Lymphoblastic Leukemia: A Real-Life Multicenter Retrospective Study in Seven AIEOP (Associazione Italiana di Ematologia e Oncologia Pediatrica) Centers | Wrong outcome |
| Bitar (2014) | Population-based analysis of invasive fungal infections, France, 2001-2010 | Wrong population |
| Boldis (2023) | High incidence of Pneumocystis jirovecii pneumonia in oncological patients: a 19-year study | Wrong outcome |
| Bosh’ian (2006) | Antibodies to the infective agents of opportunistic infections in blood of patients with hemoblastosis complicated with pneumonia | Wrong population |
| Bui (2018) | Initial results of the treatment for high risk neuroblastoma according to the siopen protocol in vietnam national children's hospital | Wrong outcome |
| Calero-Bernal (2016) | Intermittent Courses of Corticosteroids Also Present a Risk for Pneumocystis Pneumonia in Non-HIV Patients | Wrong population |
| Camelo (2021) | A Tale of 2 Pneumos: The Impact of Human Immunodeficiency Virus Exposure or Infection Status on Pediatric Nasopharyngeal Carriage of Streptococcus pneumoniae and Pneumocystis jiroveci: A Nested Case Control Analysis From the Pneumonia Etiology Research In Child Health Study | Wrong population |
| Camitta (1989) | Bone marrow transplantation for children with severe aplastic anemia: use of donors other than HLA-identical siblings | Wrong outcome |
| Cangir (1990) | Ewing's sarcoma metastatic at diagnosis. Results and comparisons of two intergroup Ewing's sarcoma studies | Wrong outcome |
| Castagnola (2008) | Incidence of bacteremias and invasive mycoses in children undergoing allogeneic hematopoietic stem cell transplantation: a single center experience | Wrong outcome |
| Chaudhary (1977) | Percutaneous transthoracic needle aspiration of the lung. Diagnosing Pneumocystis carinii pneumonitis | Wrong outcome |
| Cheung (1994) | An outbreak of Pneumocystis carinii pneumonia in children with malignancy | Wrong outcome |
| de la Morena (2017) | Long-term outcomes of 176 patients with X-linked hyper-IgM syndrome treated with or without hematopoietic cell transplantation | Wrong outcome |
| Dincol (1998) | A comparison of imipenem monotherapy versus cefoperazone/sulbactam plus amikacin combination treatment in febrile neutropenic cancer patients | Wrong population |
| Diri (2015) | Retrospective review of intravenous pentamidine for Pneumocystis pneumonia prophylaxis in patients undergoing hematopoietic stem cell transplantation | Wrong population |
| Driscoll (2011) | Allogeneic transplantation for patients with high-risk or refractory neuroblastoma | Wrong outcome |
| Du Plessis (2016) | Laboratory-based surveillance of Pneumocystis jirovecii pneumonia in South Africa, 2006-2010i | Wrong outcome |
| Essa (2022) | Outcomes of blinatumomab based therapy in children with relapsed, persistent, or refractory acute lymphoblastic leukemia: a multicenter study focusing on predictors of response and post-treatment immunoglobulin production | Wrong outcome |
| Evernden (2020) | High incidence of Pneumocystis jirovecii pneumonia in allogeneic hematopoietic cell transplant recipients in the modern era | Wrong population |
| Falkson (1971) | Pneumocystis carinii pneumonia in acute lymphatic leukaemia | Wrong study design |
| Fleury (1985) | Cell population obtained by bronchoalveolar lavage in Pneumocystis carinii pneumonitis | Wrong population |
| Furukawa (1977) | [Eight cases of Pneumocystis carinii pneumonitis associated with hematological malignancies (author's transl)] | Wrong study design |
| Garaventa (1995) | Fatal pneumopathy in children after bone marrow transplantation--report from the Italian Registry | Wrong outcome |
| Garaventa (1995) | Fatal pneumopathy in children after bone marrow transplantation report from the Italian Registry | Wrong outcome |
| Gennery (2004) | Treatment of CD40 ligand deficiency by hematopoietic stem cell transplantation: a survey of the European experience, 1993-2002 | Wrong outcome |
| Glazer (1998) | Use of fiberoptic bronchoscopy in bone marrow transplant recipients | Wrong population |
| Goetz (1974) | Pneumocystis carnii pneumonia | Wrong outcome |
| Goussetis (2015) | Infectious complications following allogeneic stem cell transplantation by using anti-thymocyte globulin-based myeloablative conditioning regimens in children with hemoglobinopathies | Wrong outcome |
| Haddock (1982) | Pneumocystis carinii pneumonia at North Carolina Memorial Hospital | Wrong outcome |
| Hall (1987) | A critical review of the use of open lung biopsy in the management of the oncologic patient with acute pulmonary infiltrates | Wrong outcome |
| Hamidieh (2013) | Hematopoietic stem cell transplantation with a reduced-intensity conditioning regimen in pediatric patients with Griscelli syndrome type 2 | Wrong outcome |
| Han (2014) | Effect of umbilical cord MSC infusion on the pulmonary infection in haploidentical hematopoietic stem cell transplantation | Wrong population |
| Hargrave (2001) | Progressive reduction in treatment-related deaths in Medical Research Council childhood lymphoblastic leukaemia trials from 1980 to 1997 (UKALL VIII, X and XI) | Wrong outcome |
| Hughes (1976) | Infections during continuous complete remission of acute lymphocytic leukemia: during and after anticancer therapy | Wrong outcome |
| Huh (1999) | Secondary cardiac tumor in children | Wrong outcome |
| Inagaki (2019) | Pneumocystis Infection in Children National Trends and Characteristics in the United States, 1997-2012 | Wrong outcome |
| Javier (1976) | [Neonatal leukemia. Report of seven cases (author's transl)] | Wrong outcome |
| Johnson (1968) | Pneumocystis carinii pneumonia in children with cancer | Unavailable |
| Johnson (1969) | Pneumocystis carinii pneumonia in children with cancer | Wrong outcome |
| Johnson (1970) | Pneumocystic carinii pneumonia in children with cancer. Diagnosis and treatment | Wrong outcome |
| Kay (1983) | Infections after bone marrow transplantation using cyclosporine | Wrong population |
| Khawaja (2001) | Bone marrow transplantation for CD40 ligand deficiency: a single centre experience | Wrong population |
| Kim (2010) | Early engraftment of G-CSF-primed unmanipulated allogeneic bone marrow transplantation in paediatric patients | Wrong outcome |
| Kim (2012) | Non-bacterial infections in Asian patients treated with alemtuzumab: a retrospective study of the Asian Lymphoma Study Group | Wrong population |
| Klemm (1978) | [Interstitial pneumonia in children with acute lymphoblastic leukemia. prophylaxis with pentamidine during induction chemotherapy (author's transl)] | Wrong outcome |
| Kobayashi (1976) | Pneumocystis carinii pneumonia in children with acute leukemia | Wrong study design |
| Kobayashi (2008) | The clinical feature of invasive fungal infection in pediatric patients with hematologic and malignant diseases: A 10-year analysis at a single institution at Japan | Wrong outcome |
| Kobayashi (2018) | Risk Factors for Invasive Fungal Infection in Children and Adolescents with Hematologic and Malignant Diseases: A 10-year Analysis in a Single Institute in Japan | Wrong outcome |
| Kostov (2000) | Pneumocystis carinii (PC) infection in immunocompromised patients | Unavailable |
| Kruger (2005) | Haemopoietic cell transplantation of patients with a history of deep or invasive fungal infection during prophylaxis with liposomal amphotericin B | Wrong population |
| Lampert (1977) | [Combination chemotherapy and cranial irradiation in 530 children with acute lymphoblastic leukaemia (author's transl)] | Wrong outcome |
| Le Tan Vinh (1954) | Pneumocystis pneumonia; relation to plasmocytic interstitial pneumonia | Wrong study design |
| Lee (2011) | Improved survival in multisystem langerhans cell histiocytosis with PVMC regimen | Wrong outcome |
| Lim (1974) | Direct fluorescent-antibody method for the diagnosis of Pneumocystis carinii pneumonitis from sputa or tracheal aspirates from humans | Wrong outcome |
| Link (1990) | Initial experiences with pentamidine aerosol in prevention of Pneumocystis carinii pneumonia following bone marrow transplantation | Wrong population |
| Lipsitt (2023) | Impact of Trimethoprim/Sulfamethoxazole Prophylaxis on Engraftment in Pediatric Hematopoietic Cell Transplant Recipients | Wrong outcome |
| Logan (1995) | Acute lung disease in the immunocompromised host. Diagnostic accuracy of the chest radiograph | Wrong population |
| Luna (1972) | Pneumocystis carinii pneumonitis in cancer patients | Wrong outcome |
| Maltezou (1996) | Dapsone for Pneumocystis carinii pneumonia (PCP) prophylaxis in children undergoing bone marrow transplantation (BMT) | Unavailable |
| McMullan (2014) | Once weekly trimethoprim-sulfamethoxazole for Pneumocystis jirovecii pneumonia prophylaxis in children with cancer | Wrong study design |
| McWilliams (2015) | Positive Family History, Infection, Low Absolute Lymphocyte Count (ALC), and Absent Thymic Shadow: Diagnostic Clues for All Molecular Forms of Severe Combined Immunodeficiency (SCID) | Wrong population |
| Meila (1976) | Pneumocystis carinii pneumonia in infants | Wrong study design |
| Meyers (1979) | The value of Pneumocystis carinii antibody and antigen detection for diagnosis of Pneumocystis carinii pneumonia after marrow transplantation | Wrong outcome |
| Michalak (1976) | Autoantibodies in children with Pneumocystis carinii pneumonia | Wrong study design |
| Miot (2017) | Hematopoietic stem cell transplantation in 29 patients hemizygous for hypomorphic IKBKG/NEMO mutations | Wrong outcome |
| Morse (2007) | Does atovaquone provide effective prophylaxis for Pneumocystis pneumonia in children with leukemia? | Wrong study design |
| Nishiyama (1994) | Pneumocystis carinii pneumonia | Wrong study design |
| Nouza (1992) | Pneumocystis carinii today--40 years later | Wrong study design |
| O’Sullivan (1994) | The use of aerosolized pentamidine for prophylaxis of pneumocystis-carinii pneumonia in children with leukemia | Duplicate |
| Patterson (1966) | Pneumocystis carinii pneumonia; pentamidine therapy | Unavailable |
| Pifer (1978) | Pneumocystis carinii infection: evidence for high prevalence in normal and immunosuppressed children | Wrong study design |
| Pifer (1984) | Pneumocystis carinii: a misunderstood opportunist | Wrong study design |
| Pincus (1987) | Pneumocystis carinii pneumonia in Johannesburg | Wrong outcome |
| Place (2018) | Phase I trial of the mTOR inhibitor everolimus in combination with multi-agent chemotherapy in relapsed childhood acute lymphoblastic leukemia | Wrong outcome |
| Price (1974) | Histopathology of Pneumocystis carinii infestation and infection in malignant disease in childhood | Wrong population |
| Prosperi (1966) | The pathology of interstitial pneumonia in infants | Wrong study design |
| Prucker (2009) | Induction death and treatment-related mortality in first remission of children with acute lymphoblastic leukemia: A population-based analysis of the Austrian Berlin-Frankfurt-Munster study group | Wrong outcome |
| Pui (1994) | Prevention of Pneumocystis carinii pneumonia in children with cancer | Wrong outcome |
| Ricciardi (2017) | Infectious disease ward admission positively influences P. jiroveci pneumonia (PjP) outcome: A retrospective analysis of 116 HIV-positive and HIV-negative immunocompromised patients | Wrong population |
| Rifkind (1966) | Pneumocystis carinii pneumonia. Studies on the diagnosis and treatment | Wrong population |
| Rupprecht (1981) | Lung complications in children with leucosis | Wrong outcome |
| Sangiolo (2005) | Toxicity and efficacy of daily dapsone as Pneumocystis jiroveci prophylaxis after hematopoietic stem cell transplantation: A case-control study | Wrong population |
| Sekowska (2024) | Infections with Klebsiella pneumoniae in Children Undergoing Anticancer Therapy or Hematopoietic Cell Transplantation: A Multicenter Nationwide Study | Wrong outcome |
| Sepkowitz (1992) | Pneumocystis carinii pneumonia among patients without AIDS at a cancer hospital | Wrong population |
| Sepkowitz (1992) | Pneumocystis-carinii pneumonia among patients without aids at a cancer hospital | Duplicate |
| Shepherd (1979) | Pneumocystis carinii pneumonitis: a serological study | Wrong outcome |
| Shoji (2020) | Recent epidemiology of Pneumocystis pneumonia in Japan | Wrong population |
| Slatter (2007) | Value of bronchoalveolar lavage before haematopoietic stem cell transplantation for primary immunodeficiency or autoimmune diseases | Wrong population |
| Speich (1992) | Pneumocystis carinii pneumonia in HIV-negative immunosuppressed patients | Wrong population |
| Srinivasan (2014) | Early infections after autologous hematopoietic stem cell transplantation in children and adolescents: The St. Jude experience | Wrong outcome |
| Stahel (1989) | High-dosage chemo-radiotherapy with autologous bone marrow transfusion in malignant lymphoma: indications and personal experience | Wrong population |
| Sternberg (1955) | Interstitial plasma cell pneumonia | Wrong study design |
| Tateoka (1977) | Pathohistological studies on the lungs in patients with leukemia. Part 2. Pulmonary mycosis, Pneumocystis carinii pneumonia, pulmonary tuberculosis and others. Japanese | Unable to translate |
| Timuragaoglu Irfanoglu (2001) | The frequency of Pneumocystis carinii in patients with haematologic malignancies and pneumonia | Wrong outcome |
| Tomizawa (2015) | Favorable outcome in non-infant children with MLL-AF4-positive acute lymphoblastic leukemia: a report from the Tokyo Children's Cancer Study Group | Wrong outcome |
| Tuan (1992) | Pneumocystis carinii pneumonitis following bone marrow transplantation | Wrong outcome |
| Van Eyssen (2017) | Single-centre experience of allogeneic haemopoietic stem cell transplant in paediatric patients in Cape town, South Africa | Wrong outcome |
| Vetternranta (2000) | Pediatric marrow transplantation for acute leukemia using unrelated donors and T-replete or -depleted grafts: A case-matched analysis | Wrong outcome |
| Vogel (1968) | Pneumocystis carinii pneumonia | Wrong outcome |
| Wang (2019) | Metagenomic next-generation sequencing for mixed pulmonary infection diagnosis | Wrong population |
| Wheeler (1996) | Treatment related deaths during induction and in first remission in acute lymphoblastic leukaemia: MRC UKALL X | Wrong outcome |
| Whitehurst (2024) | A comprehensive assessment of the prolonged febrile neutropenia evaluation in pediatric oncology patients | Wrong population |
| Yoo (2004) | Infectious complications and outcomes after allogeneic hematopoietic stem cell transplantation in Korea | Wrong population |
| Young (1976) | Treatment of Pneumocystis carinii pneumonia: current status of the regimens of pentamidine isethionate and pyrimethamine-sulfadiazine | Wrong outcome |
| Yuan (2024) | Co-transplantation of umbilical cord mesenchymal stem cells and peripheral blood stem cells in children and adolescents with refractory or relapsed severe aplastic anemia | Wrong outcome |
| Zajac-Spychala (2019) | Infections in children with acute myeloid leukemia: increased mortality in relapsed/refractory patients | Wrong outcome |
| Zwitserloot (2012) | Importance of neutropenia for development of invasive infections at various phases of treatment for hemato-oncological diseases in children | Wrong outcome |
