## Supplemental Table S2 for "A systematic review and meta-analysis of the incidence of *Pneumocystis jirovecii* pneumonia (PJP) in children and young people with cancer, cancer-like conditions or haematopoietic stem cell transplants"

**Supplemental Table S2. Study characteristics of included studies (with full data extraction)**

References are listed at the end of the file

| **Author (year), sample size** | **Country** | **Recruitment period** | **Study design** | **Median age, y (range)** | **% Males** | **Included no prophylaxis cohort (Y/N)** | **Included prophylaxis cohort (Y/N)** | **Line(s) of prophylaxis** | **Type of prophylaxis** | **Other population details** |
| --- | --- | --- | --- | --- | --- | --- | --- | --- | --- | --- |
| **ALL only studies** | | | | | | | | | | |
| Aur (1972)^1^, N=16 | USA | 1967-1969 | Not clear | NR | 37.5% | Y^#^ | N^#^ | NR | NR | All in haematologic relapse |
| Hughes (1975)^2^, N=180 | USA | 1972-1974 | RCT | 5  (0.3-19) | 57% | Y^#^ | N^#^ | NA | NA | 161 Caucasian, 18 Black, 1 Asian |
| Iyer (1976)^3^, N=148 | USA | 1967-1975 | RC | NR | NR | Y^#^ | N^#^ | NA | NA |  |
| Iacuone (1977)^4^, N=70 | USA | 1971-1976 | RC | NR | NR | Y^#^ | N^#^ | NA | NA |  |
| Aur (1978)^5^, N=282 | USA | 1972-1975 | RCT | 4  (0.25-9) | 56% | Y | N | NA | NA |  |
| Siegel (1980)^6^, N=844 | USA | 1972-1975 | Post-hoc analysis of 2 RCTs | ≤16 | NR | Y^#^ | N^#^ | NA | NA |  |
| Morgan (1981)^7^, N=172 | USA | 1975-1980 | RC | NR | NR | Y | Y | First | TMP/SMX |  |
| Saarinen (1984)^8^, N=100 | Finland | 1975-1981 | RC | NR | 50% | Y | N | NA | NA |  |
| Goorin (1985)^9^, N=30^a^ | USA | 1979-1982 | RCT | 4.5  (1-16) | 67% | Y | N | NA | NA | Data is for placebo group only |
| Hughes (1987)^10^, N=167 | USA | 1984-1986 | RCT | NR | NR | N | Y | NR but assumed first | TMP/SMX |  |
| van Eys (1987)^11^, N=126 | USA | NR | RCT | ≤21 | NR | Y | Y | NR but assumed first | TPM/SMX |  |
| Silverman (1997)^12^, N=23 | USA, Canada, Puerto Rico | 1985-1995 | RC | 7 months (23 days-11 months) | 61% | Y^#^ | N^#^ | NA | NA | 13% with CNS leukaemic disease at diagnosis |
| Postma (1998)^13^*, N=185 | Netherlands | NR | NR | NR | NR | N | Y | NR | TMP/SMX |  |
| Poulsen (2001)^14^, N=71 | Denmark | 1992-1997 | RC | 3.8 (1.2-14.8) | 63% | Y | N | NA | NA | All non-B cell ALL. Maintenance treatment only. Did receive TMP/SMX during induction treatment |
| Schroder (2001)^15^, N=190 | Denmark | 1992-1997 | RC | 4 (NR) | NR | Y | Y | NR but assumed first | TMP/SMX |  |
| Agger (2002)^16^, N=204 | Denmark | 1992-1997 | RC | Mean: 4 | 56% | Y | Y | NR but assumed first | TMP/SMX |  |
| Salzer (2009)^17^, N=115 | USA | 1996-2000 | RC | All <12 months | NR | N | Y | First or second | TMP/SMX or pentamidine | Treated on CCG 1953 protocol |
| Levinsen (2012)^18^, N=447 | Denmark, Finland, Iceland, Norway, Sweden | 1992-1996 | RC | NR | 53% | Y | Y | NR but assumed first | TMP/SMX |  |
| O’Hare (2012)^19,20^*, N=165 | UK | NR | RC | NR | NR | N | Y | First | TMP/SMX or dapsone |  |
| Salzer (2012)^21^, N=209 | USA | 1996-2006 | Single-arm clinical trial | All <12 months | NR | N | Y | First or second | TMP/SMX or pentamidine | Treated on CCG P9407 protocol |
| Kahn (2015)^22^*, N=730 | USA | 2005-2011 | RC | NR | NR | N^#^ | Y^#^ | NR | NR | Treated on Protocol 05-001 |
| Barnbrock (2024)^23^, N=6,136 | Germany, Austria, Switzerland, Italy, Israel, Czech Republic, Australia | 2010-2017 | RCT (phase 3) | 5.2  (1-18) | 58% | NR | Y | NR | NR | Prophylaxis given at the discretion of responsible physician |
| **Other cancer studies** | | | | | | | | | | |
| Perera (1970)^24^, N=827 | USA | 1962-1969 | RC | NR | NR | Y^#^ | N^#^ | NR | NR | 6 subgroups of cancer (largest being ALL, 45%) |
| Hughes (1973)^25^, N=872 | USA | 1962-1971 | RC | NR | NR | Y^#^ | N^#^ | NA | NA | 5 subgroups of cancers (largest being leukaemia, 78.4%) |
| Cangir (1976)^26^, N=102 | USA | 1973-1975 | Single-arm clinical trial | NR | NR | Y^#^ | N^#^ | NA | NA | All relapsed or metastatic solid tumours |
| Chusid (1978)^27^, N=291 | USA | 1965-1977 | RC | NR | NR | Y^#^ | N^#^ | NA | NA | 3 subgroups of cancers (largest being ALL, 79%). |
| Wolff (1978)^28^, N=151^a^ | USA | 1975-1976 | RC and RCT | NR | NR | Y^#^ | N^#^ | NA | NA | 2 subgroups of cancer (largest being ALL, 70%) |
| Aur (1979)^29^, N=25 | USA | 1968-1975 | RC | 10 (3-17) | 60% | NR | NR | NR | NR | Patients with NHL. Paper reported in Spanish |
| Harris (1980)^30^, N=258 | USA | 1977-1979 | RC | NR | NR | Y | Y | NR | TMP/SMX | 2 subgroups of cancer (largest being solid tumours, 52%) |
| Wilber (1980)^31^, N=823^a^ | USA | 1970-1975 | RC | NR | NR | Y | N | NA | NA | 5 subgroups of cancer (largest being Wilm’s tumour 32%) |
| Freeman (1988)^32^, N=34 | USA | 1984-1986 | Single-arm clinical trial | 86.5 (37-216) months | 56% | Y^#^ | N^#^ | NA | NA | Patients with brain stem tumours receiving hyper-fractionated radiotherapy |
| Green (1989)^33^, N=153 | USA | 1979-1986 | RCT | NR | NR | Y^#^ | N^#^ | NA | NA | Patients with Wilm’s tumour |
| Wollner (1993)^34^, N=40 | USA | 1971-1986 | RC | 10 (NR) | 65% | Y^#^ | N^#^ | NA | NA | Patients with primary peripheral nodal lymphoma (taken from a larger NHL cohort) |
| Mustafa (1994)^35^, N=60 | USA | 1989-1993 | Not clear | 12 (3-19) | 48% | N | Y | First and second | Pentamidine | 4 subgroups of cancers (largest being ALL, 77%) |
| Lindemulder (2007)^36^, N=482 | USA | 1993-2002 | RC | NR | NR | N | Y | NR | TMP/SMX | 2 subgroups of cancers (largest being leukaemia, 72%) |
| Ohata (2009)^37^, N=168 | Japan | 2003-2007 | RC | NR | NR | N | Y | First or second | TMP/SMX or pentamidine | 5 subgroups of cancers (largest being leukaemia, 37%) |
| Kim (2010)^38,39^, N=12 | Korea | 2004-2008 | PC | 8 (3-16) | 41% | N | Y | NR | TMP/SMX or pentamidine | Patients with diffuse pontine glioma |
| Esbenshade (2011)^40^, N=167 | USA | 1994-2009 | RC | NR | 60% | N | Y | NR | Dapsone | 7 subgroups of cancer (largest being pre-B cell ALL, 63%). 23% had HSCT |
| Ruggiero (2013)^41^, N=18 | Italy | 2006-2009 | Single-arm clinical trial | 11.2 (7.6-17.1) | 44% | N | Y | NR | TPM/SMX | 4 subgroups of cancers (largest being brainstem glioma, 61%). All relapsed/ refractory disease |
| Caselli (2014)^42^, N=2,466 | Italy | 2009-2011 | PC | NR | NR | N | Y | First | TMP/SMX | 2 subgroups of cancers (largest being leukaemia/ lymphoma, 56%) |
| Guest (2014)^43^*, N=33,067 | USA | 2004-2009 | RC | NR | 54% | N^#^ | Y^#^ | NR | NR | 6 subgroups of cancers (largest being non-CNS solid tumours, 36%) |
| Yanik (2015)^44^, N=50 | USA, Canada | 2005-2009 | Single-arm clinical trial | 6 (1.1-22.8) | 76% | N | Y | NR | Per institution guidance | Patients with neuroblastoma |
| Solodokin (2016)^45^, N=121 | USA | 2009-2014 | RC | 5.5 (IQR 3-13) | 61% | N | Y | First and second | Pentamidine | 16 had HSCT at prior institutions |
| Fischer (2017)^46^, N=23 | USA | 2006-2014 | RC | NR | NR | Y | N | NA | NA | Patients with retinoblastoma |
| Bustamante (2018)^47^*, N=52 | USA | 2016-2017 | RC | NR | NR | N | Y | NR | TMP/SMX | 2 subgroups of cancers (largest being leukaemia/ lymphoma, 69%) |
| Quinn (2018)^48^, N=525^b^ | USA | 2007-2014 | RC | 8 (0.08-24) | 56% | N | Y | Second | Pentamidine | 2 subgroups of cancer (largest being solid tumours/neuro-oncology, 66%) |
| Awad (2021)^49^, N=322 | USA | 2013-2017 | RC | Mean (SD): 11.3 (5.2) | 55% | N | Y | NR but assumed first | TMP/SMX | 4 subgroups of cancers (largest being ALL, 78%) |
| Meazza (2021)^50^, N=3,909 | Italy | 2009-2018 | RC | NR | NR | Y | Y | NR | TMP/SMX | 12 subgroups of cancers (largest being CNS tumours, 24%) |
| Kato (2024)^51^, N=166 | Japan | 2018-2023 | RC | NR | NR | N | Y | NR | TMP/SMX | 2 subgroups of cancers eligible (largest being haematological cancers, 58%) |
| **HSCT studies** | | | | | | | | | | |
| Solberg (1971)^52^, N=9 | USA | 1968-1970 | Case series | 7  (0.41-15) | 67% | Y^#^ | N^#^ | NA | NA | Only allogeneic HSCTs. Mostly non-malignancies (78%) |
| Meyers (1982)^53^, N=281 | USA | 1969-1979 | RC | NR  (0-19) | NR | Y^#^ | N^#^ | NA | NA | Only allogeneic HSCTs |
| Valteau (1988)^54^, N=165 | France | 1979-1986 | RC | Mean (SD): 82.1 (52.1) months | 69% | Y^#^ | N^#^ | NA | NA | Only autologous HSCTs. Mixed disease (largest cohort being neuroblastoma 35%) |
| Souza (1999)^55^, N=139 | USA | 1993-1996 | RC | ≤21 | NR | N | Y | First or second | TMP/SMX or dapsone | 100% allogeneic HSCTs |
| Kim (2008)^56^, N=106 | USA | 2001-2006 | RC | NR | NR | N | Y | Second | Pentamidine | Mostly allogeneic HSCTs (81%). 9 patients with aplastic anaemia included |
| O’Connor (2009)^57^*, N=111 HSCTs | USA | 2001-2007 | RC | NR | NR | N | Y | NR | Dapsone |  |
| Drillon (2010)^58^*, N=320 | USA | 1996-2007 | RC | NR | NR | N | Y | First | Pentamidine | Only allogeneic HSCTs. 69% patients with malignancies |
| DeMasi (2013)^59,60^, N=137 (167 HSCTs) | USA | 2005-2011 | RC | Mean: 7.8 | 57% | N | Y | First or second | Pentamidine | Mostly autologous HSCTs (68%). Mixed disease (largest being neuroblastoma, 19%) |
| Qualter (2014)^61^, N=193 (235 HSCTs) | USA | NR | RC | Mean (SD): 8.6 (6.3) | 60% | N | Y | NR | TMP/SMX or pentamidine | Mostly allogeneic HSCTs (80%). Mostly malignant diseases (73%) |
| Clark (2015)^62,63^, N=287 | USA | 2010-2013 | RC | 5  (0.2-32) | NR | N | Y | Second or more | Pentamidine |  |
| Curi (2016)^64,65^*, N=142 (142 HSCTs) | USA | 2007-2012 | RC | 7.6 (0.08-17.87) | 54% | N | Y | First | Pentamidine | Only allogeneic HSCTs. Mostly haematological malignancies (62%) |
| Levy (2016)^66,67^, N=111 (141 HSCTs) | USA | 2006-2013 | RC | 4.5 (NR) | 59% | N | Y | Second | Pentamidine | Mostly allogeneic HSCTs (81%).8 subgroups of cancer (largest being leukaemias, 37%) |
| Quinn (2018)^48^, N=228^b^ (229 HSCTs) | USA | 2007-2014 | RC | NR | NR | N | Y | Second | Pentamidine |  |
| Ardura (2019)^68^*, N=11,056 | USA | 2007-2017 | RC | NR | NR | N^#^ | Y^#^ | NR | NR |  |
| Czyzewski (2019)^69^, N=650 (650 HSCTs) | Poland | 2012-2015 | RC | NR | NR | N | Y | NR but assumed first | TMP/SMX | Mostly allogeneic HSCTs (77%). Mixed disease (largest cohort being ALL, 23%) |
| Ambrosino (2020)^70^*, N=319 | USA | 2013-2018 | RC | NR | NR | N | Y | NR | Pentamidine |  |
| Mandava (2020)^71^*, N=114 (118 HSCTs) | USA | 2007-2019 | RC | NR | NR | N | Y | NR but assumed first for TMP/SMX | TMP/SMX, pentamidine, atovaquone | Only allogeneic HSCTs |
| Lee (2023)^72^, N=240 | South Korea | 2009-2018 | RC | 11.7 (IQR 6.5-15) | 62% | N^#^ | Y^#^ | NR | NR | Only allogeneic HSCTs. 4 subgroups of haematological malignancies (largest cohort being ALL, 45%) |

* Conference abstract/letter to editor. ^#^ Prophylaxis details were not reported for these studies, so judgments were made by a clinician on the likelihood of prophylaxis use based on study year/recruitment period and population characteristics.

^a^ These studies include a cohort of <100 ALL patients receiving prophylaxis that have been reported in the basic data extraction table

^b^ Quinn (2018) is presented twice in the table due to separate extractable cohorts - this is the same study.

ALL = acute lymphoblastic leukaemia; F = female; HSCT = haematopoietic stem cell transplant; IQR = interquartile range; M =male; N = no; NA = not applicable; NHL = non-Hodgkins lymphoma; NR = not reported; PC = prospective cohort; RC = retrospective cohort; RCT = randomised control trial; SD = standard deviation; TMP/SMX = trimethoprim-sulfamethoxazole; UK = United Kingdom; USA = United States of America; Y = yes
