## Supplemental Table S3 for "A systematic review and meta-analysis of the incidence of *Pneumocystis jirovecii* pneumonia (PJP) in children and young people with cancer, cancer-like conditions or haematopoietic stem cell transplants"

**SUPPLEMENTAL TABLE S3 PJP Results for HSCT studies**

| **Author (year)** | **Prophylaxis (% of patients)** | **Sample size** | **Total N of PJP cases^a^** | **Number of PJP cases** | | | **N of PJP patients died (%)** | **Timing of PJP diagnosis** | **Details of PJP cases** |
| --- | --- | --- | --- | --- | --- | --- | --- | --- | --- |
|  |  |  |  | **N CONF** | **N SUSP** | **N UNC** |  |  |  |
| **HSCT studies** | | | | | | | | | |
| Solberg (1971)^1^ | NR but assumed no^#^ | 9 | 2 | NR | NR | 2 | 2 (100) | NR | Both male, under two years old with sex-linked lymphopenic hypogammaglobulinaemia receiving allogeneic HSCT |
| Meyers (1982)^2^ | NR but assumed no^#^ | 281 | 18 | 18 | NR | - | NR | NR | All cases received allogeneic HSCT |
| Valteau (1988)^3^ | NR but assumed no^#^ | 165 | 1 | NR | NR | 1 | 0 | Day +192 post-HSCT | Received autologous HSCT |
| Souza (1999)^4^ | 160mg/kg TMP + 800mg/kg SMX (91), 50mg dapsone (9) | 139 | 0 | - | - | - | NA | NA |  |
| Kim (2008)^5^ | 4mg/kg IV pentamidine (100) | 106 | 2 | 2 | NR | - | NR | NR |  |
| O’Connor (2009)*^6^ | Dapsone (100) | 111 (N of HSCTs) | 0 | - | - | - | NA | NA |  |
| Drillon (2010)*^7^ | 4mg/kg IV pentamidine (100) | 320 | 0 | - | - | - | NA | NA |  |
| DeMasi (2013)^8,9^ | 4mg/kg IV pentamidine (100) | 137 (167 HSCTs) | 0 | - | - | - | NA | NA |  |
| Qualter (2014)^10^ | 5mg/kg TMP/SMX or 4mg/kg pentamidine (100) | 193 (235 HSCTs) | 1 | 1 | NR | - | NR | NR |  |
| Clark (2015)^11,12^ | 4mg/kg IV pentamidine (100) | 287 | 1 | 1 | NR | - | NR | Day+15 post 2^nd^ HSCT |  |
| Curi (2016)*^13,14^ | 4mg/kg IV pentamidine (100) | 142 | 0 | - | - | - | NA | NA |  |
| Levy (2016)^15,16^ | 4mg/kg IV pentamidine (100) | 111 (141 HSCTs) | 0 | - | - | - | NA | NA |  |
| Quinn (2018)^17^ | Up to 300mg per dose, aerosolised pentamidine (15) | 34 (N of HSCTs) | 0 | - | - | - | NA | NA |  |
|  | 3-4mg/kg IV pentamidine (69) | 158 (N of HSCTs) | 1 | - | 1 | - | NR | NR | Suspected case was 11-month old female with relapsed acute lymphoblastic leukaemia |
|  | Aerosolised and IV pentamidine (16) | 37 (N of HSCTs) | 0 | - | - | - | NA | NA |  |
| Ardura (2019)*^18^ | NR but assumed yes^#^ | 11,056 | 130 | NR | NR | 130 | NR | NR | 22 HSCTs were autologous, 113 HSCTs were allogeneic |
| Czyzewski (2019)^19^ | TMP/SMX (100) | 650 | 1 | NR | NR | 1 | NR | NR |  |
| Ambrosino (2020)*^20^ | Pentamidine (100) | 319 | 4 | NR | NR | 4 | NR | NR |  |
| Mandava 2020*^21^ | 2.5mg/kg TMP/SMX (78) | 90 (N of HSCTs) | 1 | NR | NR | 1 | 1 (100) | Day +198 post-HSCT | Patient with leukaemia who received allogeneic HSCT |
|  | Pentamidine (22) | 25 (N of HSCTs) | 0 | - | - | - | NA | NA |  |
|  | Atovaquone (1) | 1 | 0 | - | - | - | NA | NA |  |
| Lee (2023)^22^ | NR but assumed yes^#^ | 240 | 5 | NR | 5 | - | NR | NR | All allogeneic HSCTs |

* Conference abstract/letter to editor. ^#^ Prophylaxis details were not reported for these studies, so judgments were made by a clinician on the likelihood of prophylaxis use based on study year/recruitment period and population characteristics.

^a^ Total number of PJP cases is the number of confirmed, suspected and unclear cases combined. All %s have been rounded to whole numbers.

CONF = confirmed; HSCT = haematopoietic stem cell transplant; IV = intravenous; N = number; NA = not applicable; NR = not reported; PJP = *pneumocystis jirovecii* pneumonia; SMX = sulfamethoxazole; SUSP = suspected; TMP = trimethoprim; UNC = unclear
