## Supplemental Table S4 for "A systematic review and meta-analysis of the incidence of *Pneumocystis jirovecii* pneumonia (PJP) in children and young people with cancer, cancer-like conditions or haematopoietic stem cell transplants"

**SUPPLEMENTAL TABLE S4 PJP results for studies reporting an ALL population**

| **Author (year)** | **Prophylaxis (% of patients)** | **Sample size** | **Total N of PJP cases^a^** | **Number of PJP cases** | | | **N of PJP patients died (%)** | **Timing of PJP diagnosis** | **Details of PJP cases** |
| --- | --- | --- | --- | --- | --- | --- | --- | --- | --- |
|  |  |  |  | **N CONF** | **N SUSP** | **N UNC** |  |  |  |
| **ALL-only studies** | | | | | | | | | |
| Aur (1972)^1^ | NR but assumed no^#^ | 16 | 5 | NR | NR | 5 | 3 (60) | NR | All patients in haematological relapse |
| Hughes (1975)^2^ | NR but assumed no^#^ | 180 | 29 | NR | NR | 29 | NR | NR | Of the cases, median age 5 years, 62% male, 27 Caucasian |
| Iyer (1976)^3^ | NR but assumed no^#^ | 148 | 1 | 1 | NR | NR | NR | NR |  |
| Iacuone (1977)^4^ | NR but assumed no^#^ | 70 | 5 | 1 | 4 | - | NR | Within 3 weeks of induction therapy |  |
| Aur (1978)^5^ | None (100) | 282 | 36 | NR | NR | 36 | 14 (39) | During continuation chemotherapy | 39 PJP events in total. Patients didn’t receive prophylaxis |
| Siegel (1980)^6^ | NR but assumed no^#^ | 844 | 22 | 22 | - | - | NR | All <500 days after starting treatment | 22 PJP events reported in total, not clear how many patients this relates to |
| Morgan (1981)^7^ | TMP/SMX (33) | 57 | 1 | - | 1 | - | NR | NA |  |
|  | None (67) | 115 | 16 | 8 | 8 | - | NR | Median 73.5 days from starting therapy |  |
| Saarinen (1984)^8^ | None (100) | 100 | 7 | 7 (8 episodes of PJP) | - | - | 3 (43) | Average 3 months after diagnosis, 7 episodes during remission or maintenance, 1 episode at relapse | One patient with two episodes of PJP separated by 2.5 years |
| Goorin (1985)^9^ | None (100) | 30 | 1 | NR | NR | 1 | 1 (100) | NR |  |
| Hughes (1987)^10^ | 150mg/m^2^ TMP + 750mg/m^2^ SMX twice daily (55) | 92 | 0 | - | - | - | NA | NA |  |
|  | 150mg/m^2^ TMP + 750mg/m^2^ SMX, 3 consecutive days a week (44) | 74 | 0 | - | - | - | NA | NA |  |
|  | None (1) | 1 | 1 | 1 | NR | - | NR | 2 months after chemotherapy | Patient had an allergy to sulfanomides so didn’t receive TMP/SMX prophylaxis |
| Van Eys (1987)^11^ | 4mg/kg TMP + SMX (50), None (50) | 126 | 0 | - | - | - | NA | NA |  |
| Silverman (1997)^12^ | NR but assumed no^#^ | 23 | 5 | NR | NR | 5 | 2 (40) | All during CR (1-8 months from diagnosis) |  |
| Postma (1998)*^13^ | 18mg/kg TMP/SMX, 3 or 7 days a week (100) | 185 | 14 | NR | NR | 14 | NR | NR | All cases had discontinued TMP/SMX 1-16 months (median 4 months) prior to PJP diagnosis |
| Poulsen (2001)^14^ | None (100) | 71 | 13 | 10 | 3 | - | NR | Within 8 months (3-8) of starting maintenance treatment | Of confirmed cases, 8 male, 2 female. Of suspected cases, 2 female, 1 male |
| Schroder (2001)^15^ | 10-30mg/kg SMX + 2-6 mg/kg TMP daily (60), none (40) | 190 | 0 | - | - | - | NA | NA |  |
| Agger (2002)^16^ | 10-30mg SMX + 2-6mg/kg TMP daily (63), none (37) | 204 | 0 | - | - | - | NA | NA |  |
| Salzer (2009)^17^ | TMP/SMX or pentamidine (100) | 115 | 6 | NR | NR | 6 | NR | During: induction (n=2), intensified maintenance (n=1), routine maintenance (n=3) |  |
| Levinsen (2012)^18^ | TMP/SMX (27) | 120 | 0 | - | - | - | NA | NA |  |
|  | None (73) | 327 | 10 | NR | NR | 10 | 0 (0) | Within 7 months (n=8), at 10 months (n=1) or at 17 months (n=1) of starting maintenance therapy |  |
| O’Hare (2012)*^19,20^ | TMP/SMX (99) | 164 | 1 | - | 1 | - | NR | NR |  |
|  | Dapsone (1) | 1 | 0 | - | - | - | NA | NA |  |
| Salzer (2012)^21^ | TMP/SMX or pentamidine (100) | 209 | 3 | NR | NR | 3 | 1 (33) | NR |  |
| Kahn (2015)*^22^ | NR but assumed yes^#^ | 730 | 24 | NR | NR | 24 | NR | NR | 23/24 cases were non-Hispanic |
| Barnbrock (2024)^23^ | NR (at discretion of responsible physician) | 6,136 | 6 | 2 | 4 | - | NR | During intensive chemo (n=2), during/soon after maintenance chemo (n=4) | 5/6 cases received TMP/SMX - 4 were incompliant and 1 switched to pentamidine |
| **Mixed cancer studies including an ALL subgroup** | | | | | | | | | |
| Perera (1970)^24^ | NR but assumed no^#^ | 373 | 14 | 14 | NR | - | NR | NR |  |
| Chusid (1978)^25^ | NR but assumed no^#^ | 230 | 8 | 8 | NR | - | 2 (25) |  |  |
| Wolff (1978)^26^ | NR but assumed no^#^ | 106 | 15 | NR | NR | 15 | NR | During remission between days 40-90 from onset of chemo (1 patient NR) |  |
| Harris (1980)^27^ | 4 mg/kg/day TMP + 20mg/kg/day SMX (97) | 119 | 0 | - | - | - | NA | NA |  |
|  | None (3) | 4 | 4 | 4 | NR | - | 1 (25) | NR | The patient who died from PJP was female |
| Mustafa (1994)^28^ | 200mg/m^2^, aerosolised pentamidine (100) | 46 | 0 | - | - | - | NA | NA |  |
| Esbenshade (2011)^29^ | 2mg/kg/day Dapsone (100) | 115 | 0 | - | - | - | NA | NA |  |
| Guest (2014)*^30^ | NR but assumed yes^#^ | 7851 | 98 | NR | NR | 98 | NR | Median 347 days |  |
| Solodokin (2016)^31^ | 4mg/kg pentamidine (100) | 50 | 0 | - | - | - | NA | NA |  |
| Awad (2021)^32^ | TMP/SMX (100) | 250 | 3 | 3 | - | - | NR | NR | All confirmed cases were male |
| **Mixed cancer studies including a “leukaemia” subgroup** | | | | | | | | | |
| Hughes (1973)^33^ | NR but assumed no^#^ | 684 | 45 | 45 | NR | - | NR |  |  |
| Lindemulder (2007)^34^ | 5mg/kg/day TMP + SMX (100) | 345 | 2 | NR | NR | 2 | NR | NR | Both cases were non-compliant with prophylaxis |
| Ohata (2009)^35^ | 8mg/kg TMP + SMX, twice daily (82), 5-10mg/kg inhaled or 4mg/kg IV pentamidine (18) | 62 | 0 | - | - | - | NA | NA |  |
| **Mixed cancer studies including a “leukaemia/lymphoma” subgroup** | | | | | | | | | |
| Caselli (2014)^36^ | TMP/SMX, 3 days weekly (50) | 690 | 0 | - | - | - | NA | NA |  |
|  | TMP/SMX, 2 days weekly (18) | 244 | 2 | - | 2 | - | NR | NR | Both suspected cases failed to receive prophylaxis due to drug intolerance (n=1) or non-adherence (n=1) |
|  | TMP/SMX, 1 day weekly (32) | 439 | 0 | - | - | - | NA | NA |  |
| Bustamante (2018)*^37^ | Twice weekly TMP/SMX (100) | 36 | 0 | - | - | - | NA | NA |  |
| Quinn (2018)^38^ | Up to 300mg per dose aerosolised pentamidine (39) | 69 | 0 | - | - | - | NA | NA |  |
|  | 3-4 mg/kg IV pentamidine (50) | 88 | 1 | - | 1 | - | NR | NR | Suspected PJP was in a 1 year old male with refractory progressive T-cell ALL |
|  | Aerosolised and IV pentamidine (11) | 20 | 0 | - | - | - | NA | NA |  |

* Conference abstract/letter to editor. ^#^ Prophylaxis details were not reported for these studies, so judgments were made by a clinician on the likelihood of prophylaxis use based on study year/recruitment period and population characteristics.

^a^ Total number of PJP cases is the number of confirmed, suspected and unclear cases combined. All %s have been rounded to whole numbers.

ALL = acute lymphoblastic leukaemia; CONF = confirmed; CR = complete remission; N = number; NA = not applicable; NR = not reported; PJP = *pneumocystis jirovecii* pneumonia; SMX = sulfamethoxazole; SUSP = suspected; TMP = trimethoprim; UNC = unclear
