## Supplemental Table S5 for "A systematic review and meta-analysis of the incidence of *Pneumocystis jirovecii* pneumonia (PJP) in children and young people with cancer, cancer-like conditions or haematopoietic stem cell transplants"

**SUPPLEMENTAL TABLE S5 PJP results for studies reporting other cancer populations**

| **Author (year)** | **Prophylaxis (% of patients)** | **Sample size** | **Total N of PJP cases^a^** | **Number of PJP cases** | | | **N of PJP patients died (%)** | **Timing of PJP diagnosis** | **Details of PJP cases (where reported)** |
| --- | --- | --- | --- | --- | --- | --- | --- | --- | --- |
|  |  |  |  | **N CONF** | **N SUSP** | **N UNC** |  |  |  |
| **Solid tumours, not specified** | | | | | | | | | |
| Perera (1970)^1^ | NR but assumed no^#^ | 234 | 0 | - | - | - | NA | NA |  |
| Cangir (1976)^2^ | NR but assumed no^#^ | 102 | 5 | 5 | - | - | 4 (80) | NR |  |
| Wolff (1978)^3^ | NR but assumed no^#^ | 45 | 1 | NR | NR | 1 | NR | NR |  |
| Harris (1980)^4^ | 4 mg/kg/day TMP + 20mg/kg/day SMX (81) | 110 | 0 | - | - | - | NA | NA |  |
|  | None (19) | 25 | 1 | 1 | NR | - | 0 (0) | NR |  |
| Mustafa (1994)^5^ | 200mg/m^2^ aerosolised pentamidine (100) | 7 | 0 | - | - | - | NA | NA |  |
| Ohata (2009)^6^ | 8mg/kg TMP + SMX, twice daily (74), 5-10mg/kg inhaled or 4mg/kg IV pentamidine (26) | 43 | 0 | - | - | - | NA | NA |  |
| Caselli (2014)^7^ | TMP/SMX, 1-3 days weekly (100) | 1,093 | 0 | - | - | - | NA | NA |  |
| Guest (2014)*^8^ | NR but assumed yes^#^ | 11,742 | 23 | NR | NR | 23 | NR | NR |  |
| Solodokin (2016)^9^ | 4mg/kg pentamidine (100) | 3 | 0 | - | - | - | NA | NA |  |
| Kato (2024)^10^ | TMP/SMX (100) | 69 | 0 | - | - | - | NA | NA |  |
| **Acute myeloid leukaemia** | | | | | | | | | |
| Perera (1970)^1^ | NR but assumed no^#^ | 100 | 0 | - | - | - | NA | NA |  |
| Mustafa (1994)^5^ | 200mg/m^2^ aerosolised pentamidine (100) | 5 | 0 | - | - | - | NA | NA |  |
| Esbenshade (2011)^11^ | 2mg/kg/day dapsone (100) | 16 | 0 | - | - | - | NA | NA |  |
| Guest (2014)*^8^ | NR but assumed yes^#^ | 1,675 | 19 | NR | NR | 19 | NR | Median 70 days (IQR 0-149 days) |  |
| Solodokin (2016)^9^ | 4mg/kg pentamidine (100) | 5 | 0 | - | - | - | NA | NA |  |
| Awad (2021)^12^ | TMP/SMX (100) | 31 | 1 | 1 | NR | - | NR | NR |  |
| **Non-Hodgkins lymphoma** | | | | | | | | | |
| Perera (1970)^1^ | NR but assumed no^#^ | 14 | 2 | 2 | NR | - | NR | NR |  |
| Hughes (1973)^13^ | NR but assumed no^#^ | 26 | 1 | 1 | NR | - | NR | NR |  |
| Aur (1979)^14^ | NR but assumed no^#^ | 25 | 5 | NR | NR | 5 | 1 (20) | During maintenance therapy | 3 male, 2 female. 1 Black, 4 Caucasian. Median age (range): 6 (3-14) years |
| Mustafa (1994)^5^ | 200mg/m^2^ aerosolised pentamidine (100) | 2 | 0 | - | - | - | NA | NA |  |
| Esbenshade (2011)^11^ | 2mg/kg/day dapsone (100) | 16 | 0 | - | - | - | NA | NA |  |
| Guest (2014)*^8^ | NR but assumed yes^#^ | 2,340 | 10 | NR | NR | 10 | NR | Median 33 days (IQR 0-253 days) |  |
| **Lymphoma, not specified** | | | | | | | | | |
| Wilber (1980)^15^ | None (100) | 189 | 9 | NR | NR | 9 | NR | NR |  |
| Wollner (1993)^16^ | NR but assumed no^#^ | 40 | 2 | NR | NR | 2 | 1 (50) | During consolidation treatment |  |
| Lindemulder (2007)^17^ | 5mg/kg/day TMP + SMX (100) | 137 | 0 | - | - | - | NA | NA |  |
| Ohata (2009)^6^ | 8mg/kg TMP + SMX, twice daily (73), 5-10mg/kg inhaled or 4mg/kg IV pentamidine (27) | 11 | 0 | - | - | - | NA | NA |  |
| Solodokin (2016)^9^ | 4mg/kg pentamidine (100) | 4 | 0 | - | - | - | NA | NA |  |
| **Neuroblastoma** | | | | | | | | | |
| Perera (1970)^1^ | NR but assumed no^#^ | 58 | 2 | 2 | NR | - | NR | NR |  |
| Hughes (1973)^13^ | NR but assumed no^#^ | 78 | 3 | 3 | NR | - | NR | NR |  |
| Chusid (1978)^18^ | NR but assumed no^#^ | 39 | 2 | 2 | NR | - | 2 (100) | NR |  |
| Yanik (2015)^19^ | NR (per institution guidelines) | 50 | 0 | - | - | - | NA | NA |  |
| Meazza (2021)^20^ | 10mg/kg/day TMP + SMX (93) | 446 | 0 | - | - | - | NA | NA |  |
|  | None (7) | 35 | 0 | - | - | - | NA | NA |  |
| **Brain/CNS tumours** | | | | | | | | | |
| Ohata (2009)^6^ | 8mg/kg TMP + SMX, twice daily (83), 5-10mg/kg inhaled or 4mg/kg IV pentamidine (17) | 48 | 0 | - | - | - | NA | NA |  |
| Ruggiero (2013)^21^ | TMP/SMX (100) | 7 | 0 | - | - | - | NA | NA |  |
| Guest (2014)*^8^ | NR but assumed yes^#^ | 8,071 | 15 | NR | NR | 15 | NR | NR |  |
| Solodokin (2016)^9^ | 4mg/kg pentamidine (100) | 38 | 0 | - | - | - | NA | NA |  |
| Meazza (2021)^20^ | 10mg/kg/day TMP + SMX (62) | 587 | 0 | - | - | - | NA | NA |  |
|  | None (38) | 355 | 0 | - | - | - | NA | NA |  |
| **Hodgkin’s lymphoma** | | | | | | | | | |
| Perera (1970)^1^ | NR but assumed no^#^ | 48 | 1 | 1 | NR | - | NR | NR |  |
| Hughes (1973)^13^ | NR but assumed no^#^ | 77 | 1 | 1 | NR | - | NR | NR |  |
| Esbenshade (2011)^11^ | 2mg/kg/day dapsone (100) | 7 | 0 | - | - | - | NA | NA |  |
| Guest (2014)*^8^ | NR but assumed yes^#^ | 1,388 | 4 | NR | NR | 4 | NR | NR |  |
| Awad (2021)^12^ | TMP/SMX (100) | 20 | 0 | - | - | - | NA | NA |  |
| **Langerhans cell histiocytosis** | | | | | | | | | |
| Hughes (1973)^13^ | NR but assumed no^#^ | 7 | 1 | 1 | NR | - | NR | NR |  |
| Wilber (1980)^15^ | None (100) | 100 | 2 | NR | NR | 2 | NR | NR |  |
| Ohata (2009)^6^ | 8mg/kg TMP + SMX, twice daily (100) | 4 | 0 | - | - | - | NA | NA |  |
| Meazza (2021)^20^ | 10mg/kg/day TMP + SMX (76) | 158 | 0 | - | - | - | NA | NA |  |
|  | None (24) | 50 | 0 | - | - | - | NA | NA |  |
| **Brainstem tumours/DIPG** | | | | | | | | | |
| Freeman (1988)^22^ | NR but assumed no^#^ | 34 | 1 | NR | NR | 1 | NR | NR | PJP case associated with long-term steroid use |
| Kim (2010)^23,24^ | TMP/SMX or pentamidine (100) | 12 | 0 | - | - | - | NA | NA |  |
| Ruggiero (2013)^21^ | Twice daily TMP/SMX (100) | 11 | 0 | - | - | - | NA | NA |  |
| **Wilms tumour** | | | | | | | | | |
| Wilber (1980)^15^ | None (100) | 261 | 1 | NR | NR | 1 | NR | NR |  |
| Green (1989)^25^ | NR but assumed no^#^ | 153 | 3 | 3 | NR | - | 0 (0) | Median 21 (14-27) days post last chemotherapy | 2 right kidney tumours, 1 left kidney tumours |
| Meazza (2021)^20^ | 10mg/kg/day TMP + SMX (81) | 352 | 0 | - | - | - | NA | NA |  |
|  | None (19) | 80 | 0 | - | - | - | NA | NA |  |
| **Rhabdomyosarcoma** | | | | | | | | | |
| Chusid (1978)^18^ | NR but assumed no^#^ | 22 | 1 | 1 | NR | - | 0 (0) | NR |  |
| Wilber (1980)^15^ | None (100) | 159 | 5 | NR | NR | 5 | NR | NR |  |
| Meazza (2021)^20^ | 10mg/kg/day TMP + SMX (72) | 292 | 0 | - | - | - | NA | NA |  |
|  | None (28) | 115 | 0 | - | - | - | NA | NA |  |
| **Ewing’s sarcoma** | | | | | | | | | |
| Wilber (1980)^15^ | None (100) | 114 | 1 | NR | NR | 1 | NR | NR |  |
| Meazza (2021)^20^ | 10mg/kg/day TMP + SMX (67) | 238 | 0 | - | - | - | NA | NA |  |
|  | None (33) | 117 | 0 | - | - | - | NA | NA |  |
| **Haematological cancers, not specified** | | | | | | | | | |
| Solodokin (2016)^9^ | 4mg/kg pentamidine (100) | 12 | 0 | - | - | - | NA | NA |  |
| Kato (2024)^10^ | TMP/SMX (100) | 97 | 0 | - | - | - | NA | NA |  |
| **Sarcoma, not specified** | | | | | | | | | |
| Solodokin (2016)^9^ | 4mg/kg pentamidine (100) | 7 | 0 | - | - | - | NA | NA |  |
| Meazza (2021)^20^ | 10mg/kg/day TMP + SMX (71) | 189 | 0 | - | - | - | NA | NA |  |
|  | None (29) | 77 | 0 | - | - | - | NA | NA |  |
| **Osteosarcoma** | | | | | | | | | |
| Meazza (2021)^20^ | 10mg/kg/day TMP + SMX (65) | 216 | 0 | - | - | - | NA | NA |  |
|  | None (35) | 115 | 0 | - | - | - | NA | NA |  |
| **Germ cell tumour** | | | | | | | | | |
| Meazza (2021)^20^ | 10mg/kg/day TMP + SMX (81) | 153 | 0 | - | - | - | NA | NA |  |
|  | None (19) | 35 | 0 | - | - | - | NA | NA |  |
| **Hepatoblastoma** | | | | | | | | | |
| Meazza (2021)^20^ | 10mg/kg/day TMP + SMX (75) | 76 | 0 | - | - | - | NA | NA |  |
|  | None (25) | 25 | 0 | - | - | - | NA | NA |  |
| **Nasopharyngeal carcinoma** | | | | | | | | | |
| Meazza (2021)^20^ | 10mg/kg/day TMP + SMX (52) | 16 | 0 | - | - | - | NA | NA |  |
|  | None (48) | 15 | 0 | - | - | - | NA | NA |  |
| **Retinoblastoma** | | | | | | | | | |
| Fischer (2017)^26^ | None (100) | 274 | 0 | - | - | - | NA | NA |  |
| **Aplastic anaemia** | | | | | | | | | |
| Esbenshade (2011)^11^ | 2mg/kg/day dapsone (100) | 7 | 0 | - | - | - | NA | NA |  |
| **Other** | | | | | | | | | |
| Esbenshade (2011)^11^ | 2mg/kg/day dapsone (100) | 6 | 0 | - | - | - | NA | NA |  |
| Solodokin (2016)^9^ | 4mg/kg pentamidine (100) | 2 | 0 | - | - | - | NA | NA |  |
| Bustamante (2018)*^27^ | Twice weekly TMP/SMX (100) | 16 | 0 | - | - | - | NA | NA |  |
| Quinn (2018)^28^ | Up to 300mg per dose aerosolised pentamidine (16) | 55 | 0 | - | - | - | NA | NA |  |
|  | 3-4 mg/kg IV pentamidine (75) | 262 | 2 | - | 2 | - | NR | NR | One 4-yr male with refractory metastatic neuroblastoma; one 4-yr male with progressive, metastatic melanoma |
|  | Aerosolised and IV pentamidine (9) | 31 | 0 | - | - | - | NA | NA |  |
| Awad (2021)^12^ | TMP/SMX (100) | 21 | 0 | - | - | - | NA | NA |  |
| Meazza (2021)^20^ | 10mg/kg/day TMP + SMX (84) | 140 | 0 | - | - | - | NA | NA |  |
|  | None (16) | 27 | 0 | - | - | - | NA | NA |  |

* Conference abstract/letter to editor. ^#^ Prophylaxis details were not reported for these studies, so judgments were made by a clinician on the likelihood of prophylaxis use based on study year/recruitment period and population characteristics.

^a^ Total number of PJP cases is the number of confirmed, suspected and unclear cases combined. All %s have been rounded to whole numbers.

CNS = central nervous system; CONF = confirmed; DIPG = diffuse intrinsic pontine glioma; IQR = interquartile range; IV = intravenous; N = number; NA = not applicable; NR = not reported; PJP = *pneumocystis jirovecii* pneumonia; SUSP = suspected; TMP/SMX = trimethoprim/sulfamethoxazole; UNC = unclear
