## Supporting Methods S1 for "A systematic review and meta-analysis of the incidence of *Pneumocystis jirovecii* pneumonia (PJP) in children and young people with cancer, cancer-like conditions or haematopoietic stem cell transplants"

**Supporting Methods S1. Full Search Strategies**

**Ovid MEDLINE(R) ALL**

via Ovid <http://ovidsp.ovid.com/>

Indexed Date Range: <1946 to November 19, 2024>

Date searched: 21 November 2024

Records retrieved: 3199

1	Pneumonia, Pneumocystis/ (9393)

2	((PCP or PJP) and (infection* or pneumonia*)).ti,ab,kw. (3585)

3	((carini* or jirov?ci*) adj3 pneumonia*).ti,ab,kw. (6613)

4	(pneumocyst* adj3 pneumonia*).ti,ab,kw. (8145)

5	(interstitial plasma cell* adj2 pneumonia*).ti,ab,kw. (185)

6	or/1-5 (12836)

7	Pediatrics/ (59161)

8	Pediatric Emergency Medicine/ (551)

9	Neonatology/ (3281)

10	exp Child/ (2237366)

11	exp Infant/ (1293159)

12	Adolescent/ (2288062)

13	Young Adult/ (1082135)

14	(p?ediatric* or child* or baby or babies or infant* or infancy or toddler* or neo nat* or neo-nat* or neonat* or newborn* or new-born* or new born* or newly born* or newly-born* or preschool* or pre-school* or schoolchild* or school-child* or school child* or school-age* or school age* or underage* or under-age* or boy* or girl* or kid* or preadolescen* or pre-adolescen* or preteen* or pre-teen* or pre teen* or teen* or puberty or prepuberty or pre-puberty or pubescen* or pre-pubescen* or pre pubescen* or juvenil* or adoles* or youth*).ti,ab,kw. (3675298)

15	(young adj2 (adult* or person* or people)).ti,ab,kw. (179909)

16	or/7-15 (6130739)

17	6 and 16 (3255)

18	exp animals/ not humans.sh. (5278797)

19	17 not 18 (3204)

20	remove duplicates from 19 (3199)

**Key:**

/ or sh = indexing term (Medical Subject Heading: MeSH)

exp = exploded indexing term (MeSH)

* = truncation

? = wildcard for one additional letter

ti,ab,kw = terms in either title, abstract or keyword heading fields

adj2 = terms within two words of each other (any order)

**Embase**

via Ovid <http://ovidsp.ovid.com/>

Indexed Date Range: <1974 to 2024 November 20>

Date searched: 21 November 2024

Records retrieved: 5119

1	Pneumocystis pneumonia/ (17463)

2	((PCP or PJP) and (infection* or pneumonia*)).ti,ab,kw. (6226)

3	((carini* or jirov?ci*) adj3 pneumonia*).ti,ab,kw. (8403)

4	(pneumocyst* adj3 pneumonia*).ti,ab,kw. (10734)

5	(interstitial plasma cell* adj2 pneumonia*).ti,ab,kw. (25)

6	or/1-5 (22139)

7	pediatrics/ (97070)

8	pediatric emergency medicine/ (1749)

9	neonatology/ (6043)

10	exp child/ (3276949)

11	exp infant/ (1194977)

12	adolescent/ (1889980)

13	young adult/ (582438)

14	(p?ediatric* or child* or baby or babies or infant* or infancy or toddler* or neo nat* or neo-nat* or neonat* or newborn* or new-born* or new born* or newly born* or newly-born* or preschool* or pre-school* or schoolchild* or school-child* or school child* or school-age* or school age* or underage* or under-age* or boy* or girl* or kid* or preadolescen* or pre-adolescen* or preteen* or pre-teen* or pre teen* or teen* or puberty or prepuberty or pre-puberty or pubescen* or pre-pubescen* or pre pubescen* or juvenil* or adoles* or youth*).ti,ab,kw. (4655812)

15	(young adj2 (adult* or person* or people)).ti,ab,kw. (240157)

16	or/7-15 (6502015)

17	6 and 16 (5201)

18	Animal experiment/ not (human experiment/ or human/) (2685867)

19	17 not 18 (5170)

20	remove duplicates from 19 (5119)

**Key:**

/  = indexing term (Emtree Subject Heading)

exp = exploded indexing term (Emtree)

* = truncation

? = wildcard for one additional letter

ti,ab = terms in either title, abstract or keyword heading fields

adj2 = terms within two words of each other (any order)

**APA PsycInfo**

via Ovid <http://ovidsp.ovid.com/>

Indexed Date Range: <1806 to November 2024 Week 3>

Date searched: 21 November 2024

Records retrieved: 16

1	Pneumonia/ and (carini* or jirov?ci* or pneumocyst or interstitial plasma cell*).af. (19)

2	((PCP or PJP) and (infection* or pneumonia*)).ti,ab,id. (42)

3	((carini* or jirov?ci*) adj3 pneumonia*).ti,ab,id. (42)

4	(pneumocyst* adj3 pneumonia*).ti,ab,id. (54)

5	(interstitial plasma cell* adj2 pneumonia*).ti,ab,id. (0)

6	or/1-5 (80)

7	exp Pediatrics/ (34180)

8	child*.sh. (209614)

9	infant*.sh. (32851)

10	adolescent*.sh. (116784)

11	(p?ediatric* or child* or baby or babies or infant* or infancy or toddler* or neo nat* or neo-nat* or neonat* or newborn* or new-born* or new born* or newly born* or newly-born* or preschool* or pre-school* or schoolchild* or school-child* or school child* or school-age* or school age* or underage* or under-age* or boy* or girl* or kid* or preadolescen* or pre-adolescen* or preteen* or pre-teen* or pre teen* or teen* or puberty or prepuberty or pre-puberty or pubescen* or pre-pubescen* or pre pubescen* or juvenil* or adoles* or youth*).ti,ab,id. (1208052)

12	(young adj2 (adult* or person* or people)).ti,ab,id. (107030)

13	or/7-12 (1265365)

14	6 and 13 (16)

15	remove duplicates from 14 (16)

**Key:**

/ = indexing term (American Psychological Association's Thesaurus of Psychological Index Terms)

exp = exploded indexing term (American Psychological Association's Thesaurus of Psychological Index Terms)

* = truncation

? = wildcard for one additional letter

ti,ab,id = terms in either title, abstract or key concepts fields

adj2 = terms within two words of each other (any order)

**Maternity & Infant Care Database (MIDIRS)**

via Ovid <http://ovidsp.ovid.com/>

Indexed Date Range: <1971 to November 05, 2024>

Date searched: 21 November 2024

Records retrieved: 43

1	((PCP or PJP) and (infection* or pneumonia*)).ti,ab. (10)

2	((carini* or jirov?ci*) adj3 pneumonia*).ti,ab. (42)

3	(pneumocyst* adj3 pneumonia*).ti,ab. (48)

4	(interstitial plasma cell* adj2 pneumonia*).ti,ab. (0)

5	1 or 2 or 3 or 4 (50)

6	(p?ediatric* or child* or baby or babies or infant* or infancy or toddler* or neo nat* or neo-nat* or neonat* or newborn* or new-born* or new born* or newly born* or newly-born* or preschool* or pre-school* or schoolchild* or school-child* or school child* or school-age* or school age* or underage* or under-age* or boy* or girl* or kid* or preadolescen* or pre-adolescen* or preteen* or pre-teen* or pre teen* or teen* or puberty or prepuberty or pre-puberty or pubescen* or pre-pubescen* or pre pubescen* or juvenil* or adoles* or youth*).ti,ab. (187349)

7	(young adj2 (adult* or person* or people)).ti,ab. (1639)

8	6 or 7 (187638)

9	5 and 8 (43)

**Key:**

* = truncation

? = wildcard for one additional letter

ti,ab = terms in either title or abstract fields

adj2 = terms within two words of each other (any order)

**EB Health - KSR Evidence**

via Ovid <http://ovidsp.ovid.com/>

Indexed Date Range: <2015 to 2024 Week 45>

Date searched: 21 November 2024

Records retrieved: 13

1	((PCP or PJP) and (infection* or pneumonia*)).ti,ab,po. (58)

2	((carini* or jirov?ci*) adj3 pneumonia*).ti,ab,po. (46)

3	(pneumocyst* adj3 pneumonia*).ti,ab,po. (71)

4	(interstitial plasma cell* adj2 pneumonia*).ti,ab,po. (0)

5	1 or 2 or 3 or 4 (73)

6	(p?ediatric* or child* or baby or babies or infant* or infancy or toddler* or neo nat* or neo-nat* or neonat* or newborn* or new-born* or new born* or newly born* or newly-born* or preschool* or pre-school* or schoolchild* or school-child* or school child* or school-age* or school age* or underage* or under-age* or boy* or girl* or kid* or preadolescen* or pre-adolescen* or preteen* or pre-teen* or pre teen* or teen* or puberty or prepuberty or pre-puberty or pubescen* or pre-pubescen* or pre pubescen* or juvenil* or adoles* or youth*).ti,ab,po. (52253)

7	(young adj2 (adult* or person* or people)).ti,ab,po. (3321)

8	6 or 7 (53227)

9	5 and 8 (13)

**Key:**

* = truncation

? = wildcard for one additional letter

ti,ab,po = terms in either title, abstract or population fields

adj2 = terms within two words of each other (any order)

**EB Health - ECRI Guidelines Trust**

via Ovid <http://ovidsp.ovid.com/>

Indexed Date Range: <2018 to 2024 Week 46>

Date searched: 21 November 2024

Records retrieved: 2

1	((PCP or PJP) and (infection* or pneumonia*)).af. (1)

2	((carini* or jirov?ci*) adj3 pneumonia*).af. (1)

3	(pneumocyst* adj3 pneumonia*).af. (2)

4	(interstitial plasma cell* adj2 pneumonia*).af. (0)

5	1 or 2 or 3 or 4 (2)

**Key:**

* = truncation

? = wildcard for one additional letter

af = terms in all fields

adj2 = terms within two words of each other (any order)

**Cochrane Database of Systematic Reviews (CDSR)**

via Wiley <http://onlinelibrary.wiley.com/>

Issue 11 of 12, November 2024

Date searched: 21 November 2024

Records retrieved: 4

#1	[mh ^"Pneumonia, Pneumocystis"]	300

#2	((PCP or PJP) and (infection* or pneumonia*)):ti,ab,kw	284

#3	((carini* or jirov?ci*) NEAR/3 pneumonia*):ti,ab,kw	359

#4	(pneumocyst* NEAR/3 pneumonia*):ti,ab,kw	615

#5	("interstitial plasma" NEXT cell* NEAR/2 pneumonia*):ti,ab,kw	0

#6	{OR #1-#5}	688

#7	[mh ^Pediatrics]	952

#8	[mh ^"Pediatric Emergency Medicine"]	15

#9	[mh ^Neonatology]	63

#10	[mh Child]	83242

#11	[mh Infant]	46801

#12	[mh ^Adolescent]	138801

#13	[mh ^"Young Adult"]	99042

#14	(p*diatric* OR child* OR baby OR babies OR infant* OR infancy OR toddler* OR neo NEXT nat* OR neonat* OR newborn* OR new NEXT born* OR newly NEXT born* OR preschool* OR pre NEXT school* OR schoolchild* OR school NEXT child* OR school NEXT age* OR underage* OR under NEXT age* OR boy* OR girl* OR kid* OR preadolescen* OR pre NEXT adolescen* OR preteen* OR pre NEXT teen* OR teen* OR puberty OR prepuberty OR pre NEXT puberty OR pubescen* OR pre NEXT pubescen* OR juvenil* OR adoles* OR youth*):ti,ab,kw	454405

#15	(young NEAR/2 (adult* OR person* OR people)):ti,ab,kw	125113

#16	{OR #7-#15}	517501

#17	#6 AND #16 in Cochrane Reviews	4

**Key:**

mh ^ = unexploded subject heading (MeSH heading)

mh without ^ = exploded subject heading (MeSH heading)

* = truncation

? = wildcard for one additional letter

ti,ab = terms in title or abstract fields

NEAR/3 = terms within three words of each other

NEXT = terms are next to each other

**Science Citation Index - Expanded**

via Web of Science <https://www-webofscience-com>

Indexed Date Range: 1900-Present

Date searched: 21 November 2024

Records retrieved: 1627

9	#8 AND #5	1,627

8	#7 OR #6	3,681,423

7	TS=(young NEAR/2 (adult* or person* or people))	202,500

6	TS=(p?ediatric* or child* or baby or babies or infant* or infancy or toddler* or "neo nat*" or neo-nat* or neonat* or newborn* or new-born* or "new born*" or "newly born*" or newly-born* or preschool* or pre-school* or schoolchild* or school-child* or "school child*" or school-age* or "school age*" or underage* or under-age* or boy* or girl* or kid* or preadolescen* or pre-adolescen* or preteen* or pre-teen* or "pre teen*" or teen* or puberty or prepuberty or pre-puberty or pubescen* or pre-pubescen* or "pre pubescen*" or juvenil* or adoles* or youth*)	3,573,591

5	#4 OR #3 OR #2 OR #1	10,988

4	TS=("interstitial plasma cell*" NEAR/2 pneumonia*)	29

3	TS=(pneumocyst* NEAR/3 pneumonia*)	9,930

2	TS=((carini* or jirov?ci*) NEAR/3 pneumonia*)	9,243

1	TS=((PCP or PJP) and (infection* or pneumonia*))	3,185

**Key:**

TS= terms in either title, abstract, author keywords, and keywords plus fields

* = truncation

? = wildcard for one additional letter

NEAR/3 = terms within three words of each other

**ProQuest™ Dissertations & Theses Citation Index**

via Web of Science <https://www-webofscience-com>

Indexed Date Range: 1637-Present

Date searched: 21 November 2024

Records retrieved: 21

9	#8 AND #5	21

8	#6 OR #7	559,716

7	TS=(young NEAR/2 (adult* or person* or people))	46,080

6	TS=(p?ediatric* or child* or baby or babies or infant* or infancy or toddler* or "neo nat*" or neo-nat* or neonat* or newborn* or new-born* or "new born*" or "newly born*" or newly-born* or preschool* or pre-school* or schoolchild* or school-child* or "school child*" or school-age* or "school age*" or underage* or under-age* or boy* or girl* or kid* or preadolescen* or pre-adolescen* or preteen* or pre-teen* or "pre teen*" or teen* or puberty or prepuberty or pre-puberty or pubescen* or pre-pubescen* or "pre pubescen*" or juvenil* or adoles* or youth*)	537,682

5	#4 OR #3 OR #2 OR #1	209

4	TS=("interstitial plasma cell*" NEAR/2 pneumonia*)	0

3	TS=(pneumocyst* NEAR/3 pneumonia*)	134

2	TS=((carini* or jirov?ci*) NEAR/3 pneumonia*)	100

1	TS=((PCP or PJP) and (infection* or pneumonia*))	142

**Key:**

TS= terms in either title, abstract, author keywords, and keywords plus fields

* = truncation

? = wildcard for one additional letter

NEAR/3 = terms within three words of each other

**CINAHL Ultimate**

via EBSCO <https://web.p.ebscohost.com/>

Indexed Date Range: Inception-Present

Date searched: 21 November 2024

Records retrieved: 453

S17	S6 AND S16	(453)

S16	S7 OR S8 OR S9 OR S10 OR S11 OR S12 OR S13 OR S14 OR S15	(1,697,459)

S15	TI (young N2 (adult* or person* or people)) OR AB (young N2 (adult* or person* or people))	(76,897)

S14	TI (p?ediatric* or child* or baby or babies or infant* or infancy or toddler* or "neo nat*" or neo-nat* or neonat* or newborn* or new-born* or "new born*" or "newly born*" or newly-born* or preschool* or pre-school* or schoolchild* or school-child* or "school child*" or school-age* or "school age*" or underage* or under-age* or boy* or girl* or kid* or preadolescen* or pre-adolescen* or preteen* or pre-teen* or "pre teen*" or teen* or puberty or prepuberty or pre-puberty or pubescen* or pre-pubescen* or "pre pubescen*" or juvenil* or adoles* or youth*) OR AB (p?ediatric* or child* or baby or babies or infant* or infancy or toddler* or "neo nat*" or neo-nat* or neonat* or newborn* or new-born* or "new born*" or "newly born*" or newly-born* or preschool* or pre-school* or schoolchild* or school-child* or "school child*" or school-age* or "school age*" or underage* or under-age* or boy* or girl* or kid* or preadolescen* or pre-adolescen* or preteen* or pre-teen* or "pre teen*" or teen* or puberty or prepuberty or pre-puberty or pubescen* or pre-pubescen* or "pre pubescen*" or juvenil* or adoles* or youth*)	(1,071,571)

S13	(MH "Young Adult")	(294,317)

S12	(MH "Adolescence")	(624,213)

S11	(MH "Infant+")	(295,404)

S10	(MH "Child+")	(779,769)

S9	(MH "Neonatology")	(1,543)

S8	(MH "Pediatric Emergency Nursing")	(58)

S7	(MH "Pediatrics")	(21,764)

S6	S1 OR S2 OR S3 OR S4 OR S5	(2,130)

S5	TI ("interstitial plasma cell*" N2 pneumonia*) OR AB ("interstitial plasma cell*" N2 pneumonia*)	(0)

S4	TI (pneumocyst* N3 pneumonia*) OR AB (pneumocyst* N3 pneumonia*)	(1,325)

S3	TI ((carini* or jirov?ci*) N3 pneumonia*) OR AB ((carini* or jirov?ci*) N3 pneumonia*)	(1,038)

S2	TI ((PCP or PJP) and (infection* or pneumonia*)) OR AB ((PCP or PJP) and (infection* or pneumonia*))	(595)

S1	(MH "Pneumonia, Pneumocystis")	(1,486)

**Key:**

TI = terms in either title fields

AB = terms in abstract field

* = truncation

? = wildcard for one additional letter

N3 = terms within three words of each other

**PROSPERO**

via CRD <https://www.crd.york.ac.uk/prospero/>

Indexed Date Range: Inception-Present

Date searched: 21 November 2024

Records retrieved: 41

#1	MeSH DESCRIPTOR Pneumonia, Pneumocystis	6

#2	((PCP or PJP) and (infection* or pneumonia*))	58

#3	((carini* or jirov?ci*) adj3 pneumonia*)	41

#4	(pneumocyst* adj3 pneumonia*)	60

#5	(interstitial plasma cell* adj2 pneumonia*)	2

#6	#1 OR #2 OR #3 OR #4 OR #5	83

#7	MeSH DESCRIPTOR Pediatrics	126

#8	MeSH DESCRIPTOR Pediatric Emergency Medicine	1

#9	MeSH DESCRIPTOR Neonatology	2

#10	MeSH DESCRIPTOR Child EXPLODE ALL TREES	7039

#11	MeSH DESCRIPTOR Infant EXPLODE ALL TREES	1951

#12	MeSH DESCRIPTOR Adolescent	3346

#13	MeSH DESCRIPTOR Young Adult	449

#14	(p?ediatric* or child* or baby or babies or infant* or infancy or toddler* or neo nat* or neo-nat* or neonat* or newborn* or new-born* or new born* or newly born* or newly-born* or preschool* or pre-school* or schoolchild* or school-child* or school child* or school-age* or school age* or underage* or under-age* or boy* or girl* or kid* or preadolescen* or pre-adolescen* or preteen* or pre-teen* or pre teen* or teen* or puberty or prepuberty or pre-puberty or pubescen* or pre-pubescen* or pre pubescen* or juvenil* or adoles* or youth*)	105870

#15	(young adj2 (adult* or person* or people))	7736

#16	#7 OR #8 OR #9 OR #10 OR #11 OR #12 OR #13 OR #14 OR #15	107545

#17	#6 AND #16	41

**Key:**

MeSH DESCRIPTOR = indexing term (Medical Subject Heading: MeSH)

EXPLODE ALL TREES = exploded indexing term (MeSH)

* = truncation

? = wildcard for one additional letter

adj2 = terms within two words of each other (any order)

**International HTA Database**

via <https://database.inahta.org/>

Indexed Date Range: Inception-Present

Date searched: 21 November 2024

Records retrieved: 1

(Pneumonia, Pneumocystis)[mh] OR (((PCP or PJP) and (infection* or pneumonia*))) OR (((carini* or jiroveci*) and pneumonia*)) OR (((carini* or jirovaci*) and pneumonia*)) OR ((pneumocyst* and pneumonia*)) OR ((interstitial plasma cell* and pneumonia*)) = 1

**Key:**

[mh] = indexing term (Medical Subject Heading: MeSH)

* = truncation
