## Supporting Methods S2 for "A systematic review and meta-analysis of the incidence of *Pneumocystis jirovecii* pneumonia (PJP) in children and young people with cancer, cancer-like conditions or haematopoietic stem cell transplants"

**Supporting Methods S2. Quality assessment checklist**

Adapted JBI critical appraisal checklist for PJP prevalence studies

The tool was modified to remove the questions about survey studies as they were not expected to be included, and a question about the time period of incidence being reported was added. Only studies with at least one cohort receiving full data extraction were quality assessed.

**Questions:**

Q1. Was the sample frame and participants sampled appropriate to address the target population?

Q2. Were the study subjects and the setting described in detail?

Q3. Were valid methods used for the identification of the condition?

Q4a. Were both a numerator and denominator reported for PJP incidence proportion results?

Q4b. Were incidence data reported which allow a distinction to be made between the number of events and the number of patients?

Q4c. Did the study report, or state an aim to report (if no cases), a PJP incidence rate?

Q5. Was the duration of the incidence period reported?

**Adapted checklist responses:**

Yes

Probably yes

No

Probably no

Unclear

Not applicable
