## Supporting Methods S3 for "A systematic review and meta-analysis of the incidence of *Pneumocystis jirovecii* pneumonia (PJP) in children and young people with cancer, cancer-like conditions or haematopoietic stem cell transplants"

**Supporting Methods S3. Further meta-analysis methods**

**Synthesis methods**

For the Bayesian random effects BB model, the WinBUGS(1) code was adapted from a study by Liu et al(2), the number of iterations was 500,000 and number of thinning was 100. Convergence was assessed using trace plots. The GLMM and BB models were fitted using the logit-link function. The GLMM(3) was used as the primary meta-analysis because it is a one-stage model, that is study-specific and considers uncertainties within study variance without incorporating the continuity correction(4) (which is a fixed arbitrary value – often, 0.5) in zero cells.

The review included many studies where no PJP events occurred. The GLMM and BB approaches were considered appropriate for our analyses because they incorporate these studies’ data, whilst recognising the absence of events and both approaches do not require use of a continuity correction when estimating the pooled fixed and random effects. Like all meta-analyses, the effect sizes are weighted by the inverse of the sampling variance(5); thus, awarding greater weight to studies with larger sample sizes. For studies with zero events, the sampling variance used to calculate the incidence weight was undefined. With the GLMM and BB models the zero events are incorporated into their model structures, such that studies with less precise estimates (like zero events and small sample size) have little contribution to the pooled estimate. The GLMM and BB models do not explicitly calculate individual study effect weight like the continuity correction approach.(6-8)

Cohorts classed by authors only as “leukaemia” were included in the main ALL meta-analysis. Three studies which categorised patients only broadly as having “Leukaemia/lymphoma”(9-11) were excluded from the main analyses of ALL studies but included in sensitivity analyses. Subpopulations in studies reporting results for more than one type of disease were only included in meta-analyses if the sample size was >10.

Results were reported using both random effects and fixed effect models but given the methodological, clinical and reporting heterogeneity across the included studies in terms of study designs, populations, settings and publication years, we focused on results from the random effects analyses.

In the forest plots, an I^2^ of less than 25% was viewed as indicating results were subject to low heterogeneity, between 25% and 50% as moderate heterogeneity and over 50% as high heterogeneity.

**WINBUGS code:**

\### https://journals.plos.org/plosone/article?id=10.1371/journal.pone.0079203#pone.0079203.s001

\## beta-binomial distribution

model{

for(i in 1:N){

event[i] ~ dbin(p[i], n[i])

\## using beta-binomial approach

p[i]~ dbeta(a,b)

}

a ~ dunif(0,20000)

b ~ dunif(0,20000)

res = (a/(a+b))*1000 # scaled to 1000

}
