## Supporting Results S1 for "A systematic review and meta-analysis of the incidence of *Pneumocystis jirovecii* pneumonia (PJP) in children and young people with cancer, cancer-like conditions or haematopoietic stem cell transplants"

**Supporting Results S1. Full meta-analysis results for HSCT cohorts**

**TABLE 1 Haematopoietic Stem Cell Transplant cohort analyses**

| **Analysis subgroup** | **Number of studies** | **GLMM analysis** | | | **Beta-Binomial analysis** | |
| --- | --- | --- | --- | --- | --- | --- |
|  |  | **FE (95% CI)** | **RE (95% CI)** | **I^2^ (95% CI)** | **FE (95% CI)^1^** | **RE (95% CI)^2^** |
| 1. No prophylaxis using total number of events | 3 | 4.62 (3.03 to 6.97) | 4.27 (0.85 to 18.75) | 77.7 (28.0 to 93.1)  p=0.0112 | 7.17 (1.46 to 35.14) | 4.82 (3.00 to 7.01) |
| 2. Prophylaxis using total number of events | 15 | 1.03 (0.88 to 1.21) | 0.48 (0.22 to 1.02) | 0.0 (0.0 to 53.6)  p=0.7960 | 0.54 (0.36 to 1.12) | 0.88 (0.51 to 1.16) |
| 3. Prophylaxis using confirmed events | 10 | 0.23 (0.08 to 0.60) | 0.17 (0.03 to 0.95) | 0.0 (0.0 to 62.4)  p=0.9853 | 0.24 (0.08 to 0.70) | 0.28 (0.09 to 0.58) |
| 4. First-line prophylaxis using total number of events | 5 | 0.15 (0.04 to 0.60) | 0.15 (0.04 to 0.60) | 0.0 (0.0 to 79.2)  p=0.7429 | 0.15 (0.04 to 0.55) | 0.23 (0.05 to 0.55) |
| 5. Second-line prophylaxis using total number of events | 5 | 0.54 (0.20 to 1.42) | 0.54 (0.20 to 1.42) | 0.0 (0.0 to 79.2)  p=0.6404 | 0.54 (0.20 to 1.44) | 0.69 (0.22, 1.39) |
| **NOTE**: All results are scaled by 100%; I^2^ measures heterogeneity in 100%.  ^1^FE Beta binomial is calculated using the built-in function in R (oad)  ^2^RE is calculated via Bayesian approach using the WINBUGS code (pjpbayes.txt) via RJags. | | | | | | |

CI = confidence interval; FE = fixed effects; GLMM = generalized linear mixed model; RE = random effects

**Notes:**

Statistical analysis was performed using built-in function (GLMM, oad, Rjags) in R studio and WINBUGS. The Bayesian approach was used to perform the meta-analysis of the cumulative incidence random effect via the Beta-binomial model.

For the Bayesian approach, number of iterations was 500000 and number of thinning was 100. The convergence was assessed using the trace plots.

**Forest plots and trace plots:**

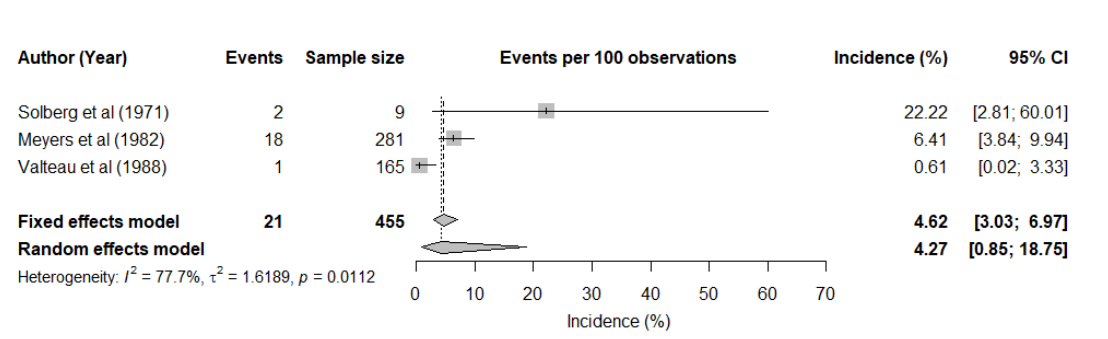

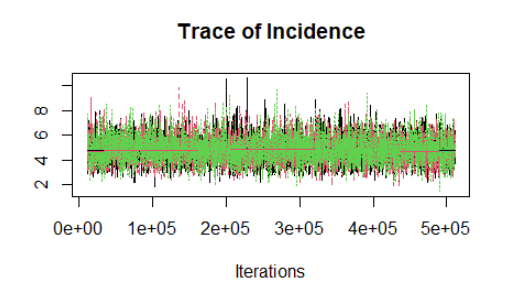

**FIGURE 1 No prophylaxis cohorts using total events**

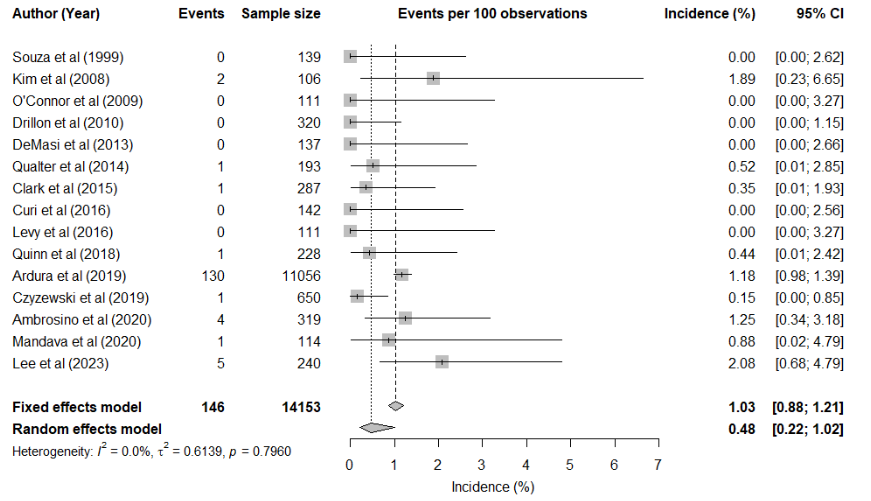

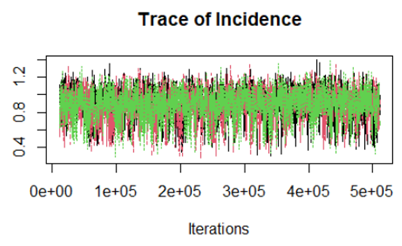

**FIGURE 2** **Prophylaxis cohorts using total events**

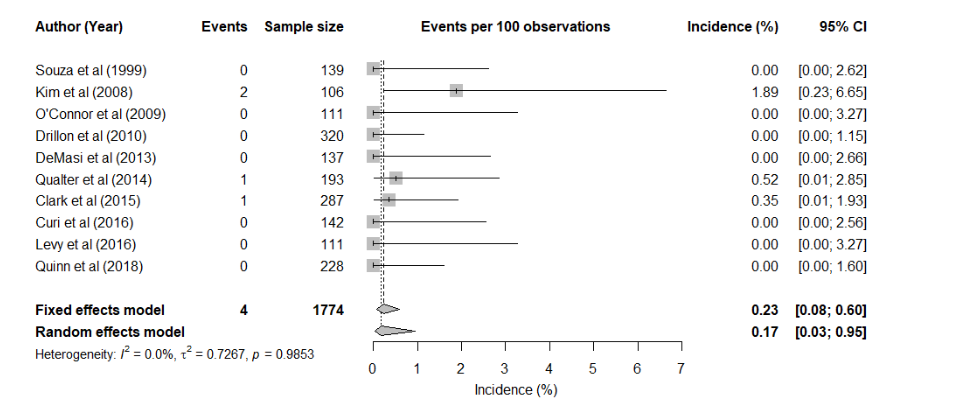

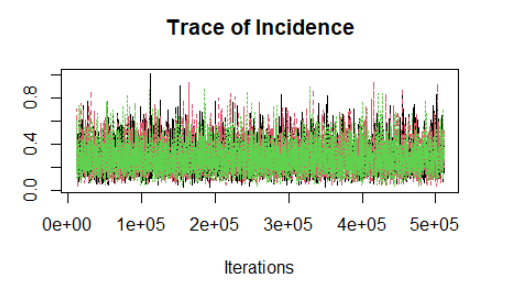

**FIGURE 3** **Prophylaxis** **cohorts using confirmed events**

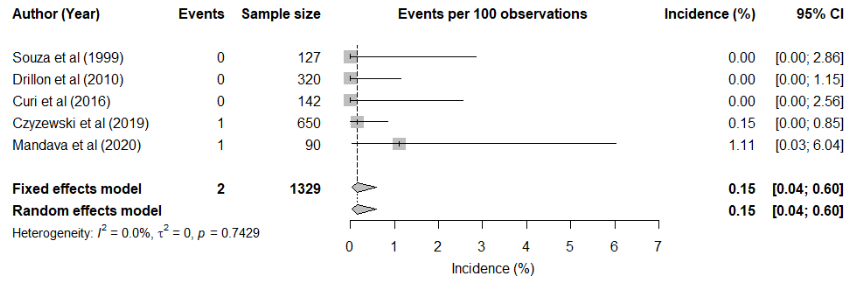

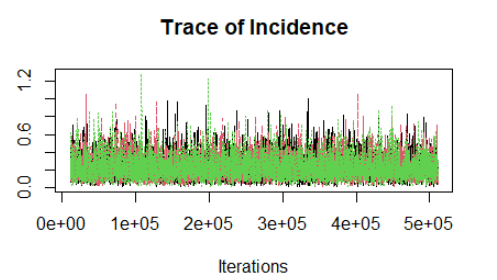

**FIGURE 4** **First-line prophylaxis cohorts using total events**

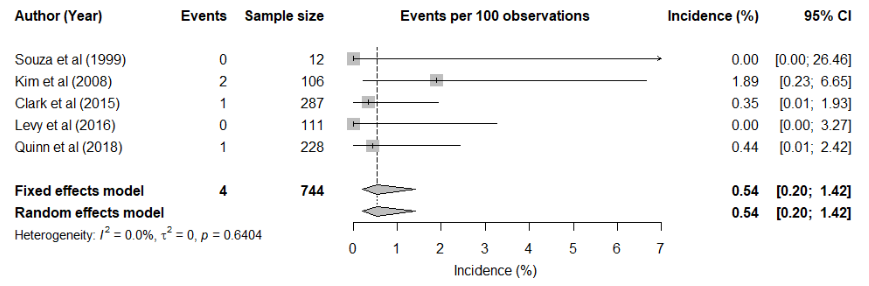

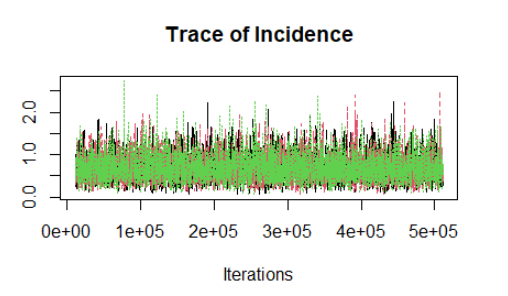

**FIGURE 5** **Second-line prophylaxis cohorts using total events**
