## Supporting Results S2 for "A systematic review and meta-analysis of the incidence of *Pneumocystis jirovecii* pneumonia (PJP) in children and young people with cancer, cancer-like conditions or haematopoietic stem cell transplants"

**Supporting Results S2. Meta-analyses: full results for the cancer cohorts**

**TABLE 1 Acute Lymphoblastic Leukaemia (ALL) analyses**

| **Population and type of event** | **Number of studies** | **GLMM analysis** | | | **Beta-Binomial analysis** | |
| --- | --- | --- | --- | --- | --- | --- |
|  |  | **FE (95% CI)** | **RE (95% CI)** | **I^2^ (95% CI)** | **FE (95% CI)^1^** | **RE (95% CI)^2^** |
| 1. No prophylaxis using total events | 19 | 6.07 (5.35, 6.87) | 5.41 (3.12, 9.21) | 85.3 (78.4, 90.0)  p<0.0001 | 8.76 (5.31, 14.44) | 7.78 (5.09, 11.83) |
| 2. No prophylaxis using confirmed events | 12 | 4.07 (3.40, 4.86) | 2.86 (1.46, 5.51) | 67.9 (41.4, 82.4)  p=0.0003 | 4.06 (2.26, 7.30) | 3.98 (2.64, 5.44) |
| 3. Prophylaxis using confirmed events^3^ | 14^3^ | 0.07 (0.03, 0.16) | 0.04 (0.003, 0.46) | 16.8 (0.0, 54.8)  p=0.27 | 0.13 (0.03, 0.58) | 0.09 (0.03, 0.20) |
| 4. Prophylaxis using confirmed events^4^  Sensitivity analysis | 17^4^ | 0.05 (0.02, 0.13) | 0.02 (0.001, 0.38) | 0.00 (0.0, 51.1)  p=0.4793 | 0.10 (0.02, 0.49) | 0.07 (0.02, 0.15) |
| 5. Prophylaxis using total events^3^ | 20^3^ | 0.93 (0.79, 1.08) | 0.49 (0.19, 1.26) | 83 (74.8, 88.5)  p<0.0001 | 1.24 (0.61, 2.50) | 1.21 (0.62, 2.17) |
| 6. Prophylaxis using total events^4^  Sensitivity analysis | 23^4^ | 0.87 (0.74, 1.01) | 0.44 (0.19, 1.04) | 82.3 (74.4, 87.7)  p<0.0001 | 1.12 (0.59, 2.14) | 1.10 (0.58, 1.90) |
| 7. First line prophylaxis using total events^4^ | 13^4^ | 0.30 (0.16, 0.58) | 0.29 (0.13, 0.66) | 0.0 (0.0, 56.6)  p=0.8812 | 0.33 (0.16, 0.68) | 0.34 (0.16, 0.59) |
| 8. Second line prophylaxis using total events^4^ | 3^4^ | 0.43 (0.06, 2.97) | 0.43 (0.06, 2.97) | 0.0 (0.0, 89.9)  p=1.0 | 0.43 (0.06, 3.06) | 0.86 (0.10, 2.35) |
| ^1^Fixed effect Beta binomial calculated using the built-in function in R (oad)  ^2^Random effects calculated via Bayesian approach using the WINBUGS code (pjpbayes.txt) via RJags.  ^3^Excluding Bustamante et al (2007), Caselli et al (2014) and Quinn et al (2018) which only reported leukaemia and lymphoma grouped together  ^4^Sensitivity analysis including Bustamante et al (2007), Caselli et al (2014) and Quinn et al (2018) which only reported leukaemia and lymphoma grouped together | | | | | | |

CI = confidence interval; FE = fixed effects; GLMM = generalized linear mixed model; RE = random effects

**ALL - Forest plots and trace plots:**

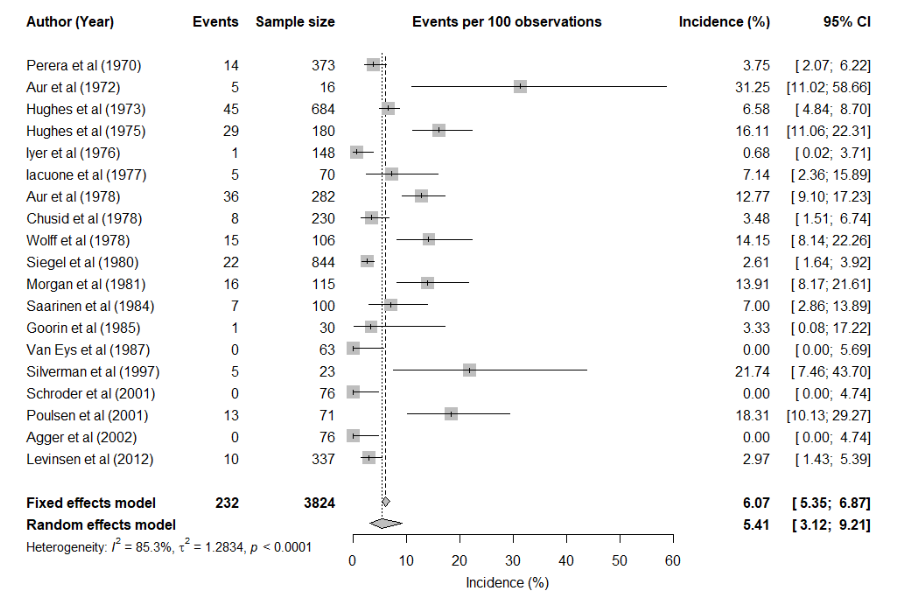

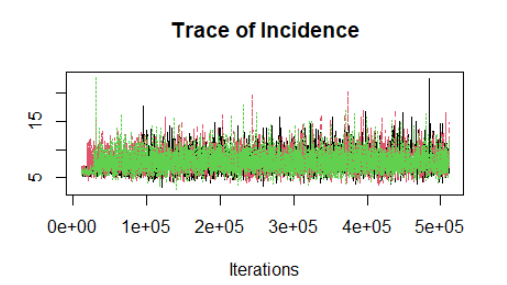

**FIGURE 1** **No prophylaxis cohorts using total events**

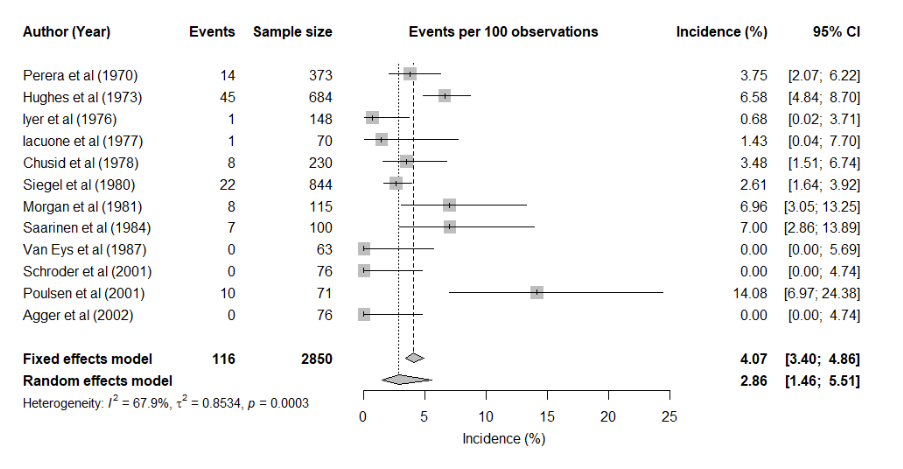

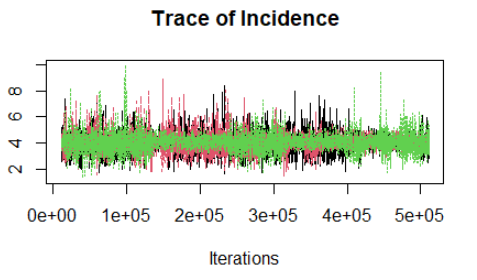

**FIGURE 2** **No prophylaxis cohorts using confirmed events**

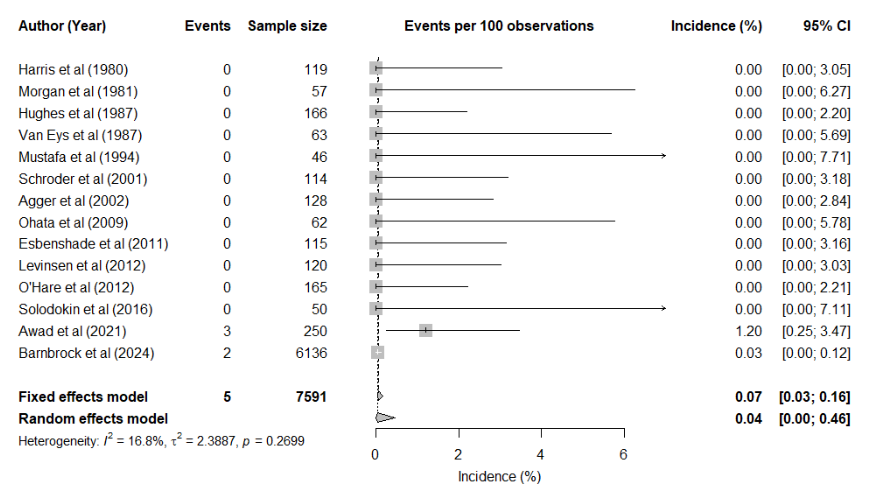

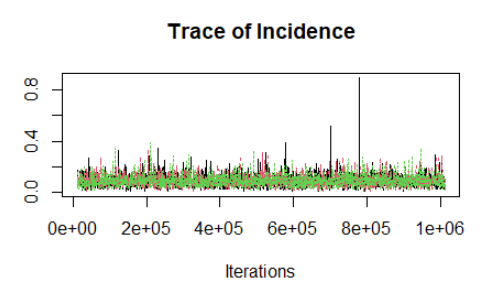

**FIGURE 3 Prophylaxis cohorts using confirmed events**

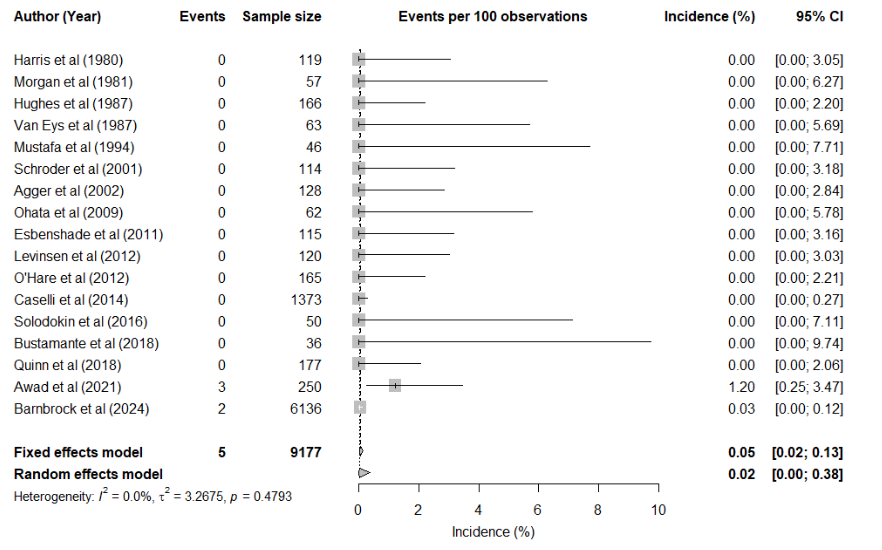

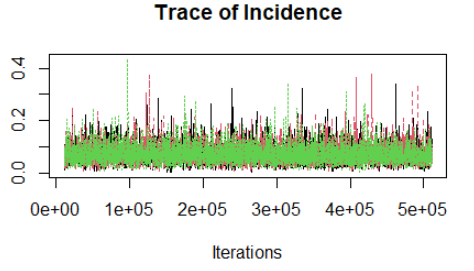

**FIGURE 4 Prophylaxis cohorts using confirmed events (sensitivity analysis)**

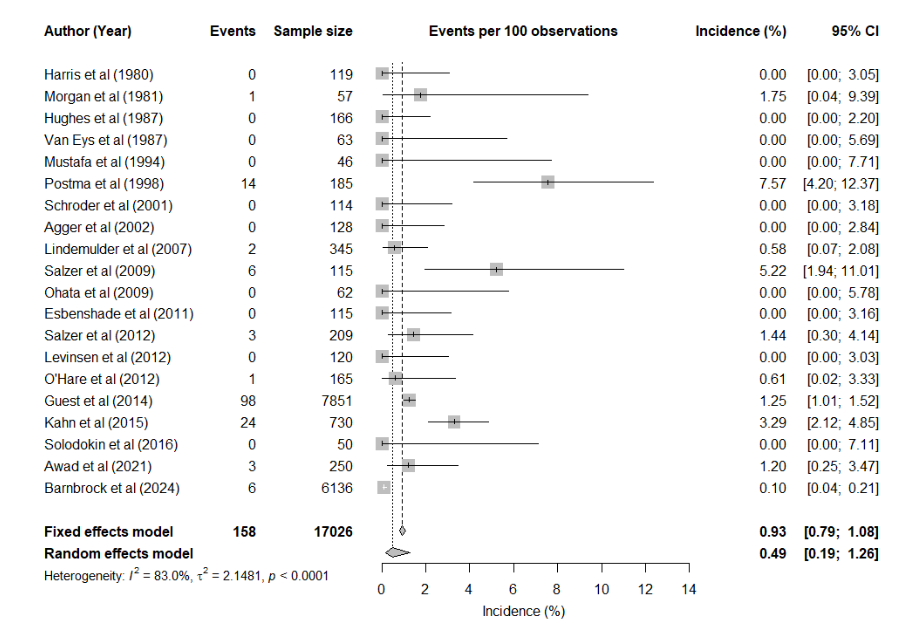

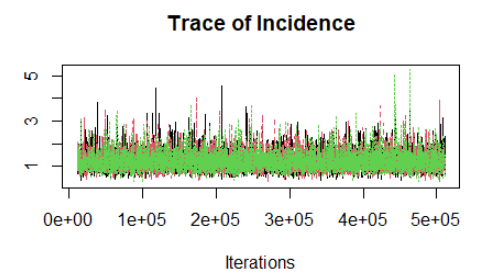

**FIGURE 5 Prophylaxis cohorts using total events**

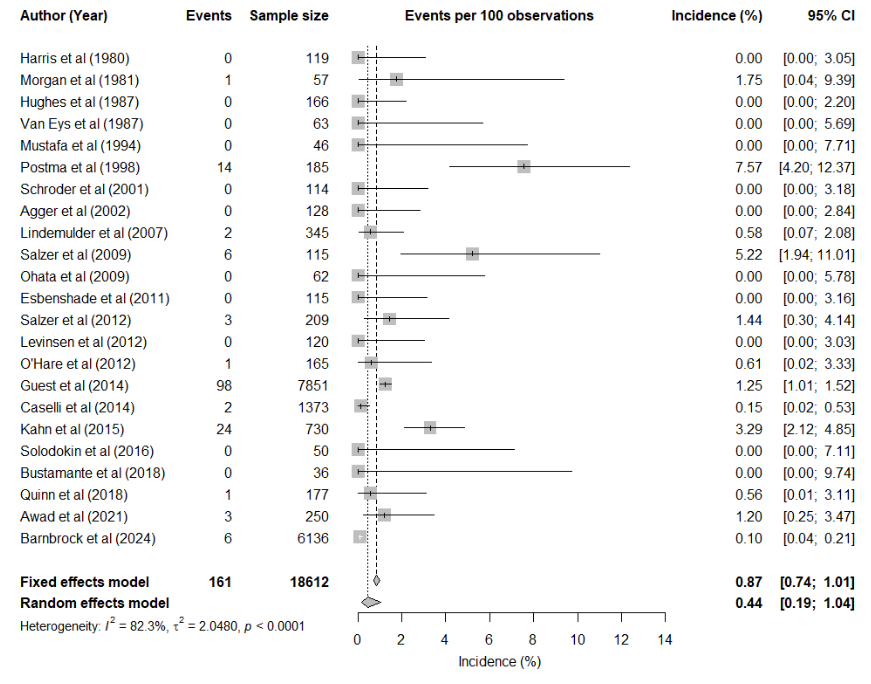

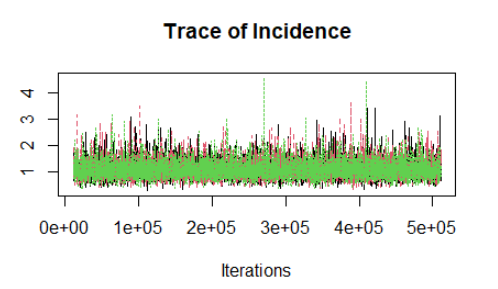

**FIGURE 6 Prophylaxis cohorts using total events (sensitivity analysis)**

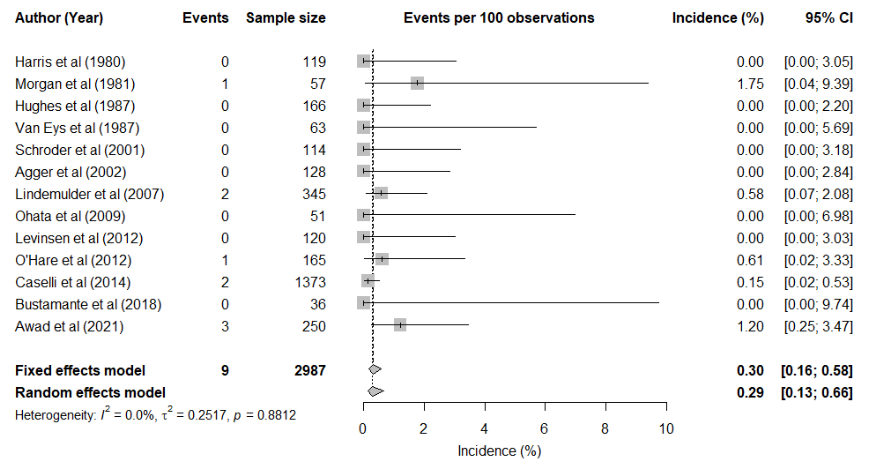

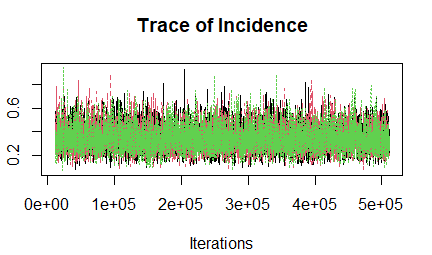

**FIGURE 7** **First-line prophylaxis cohorts using total events**

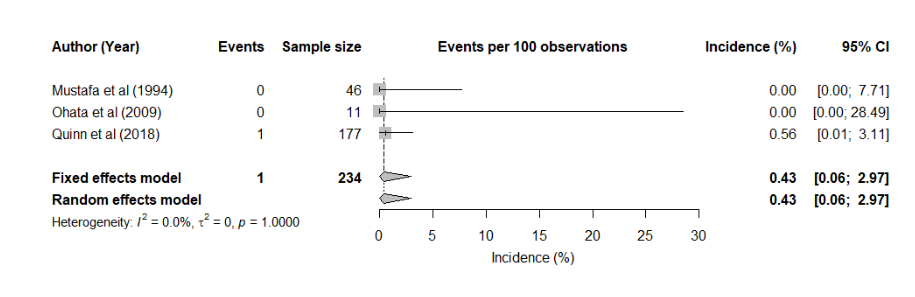

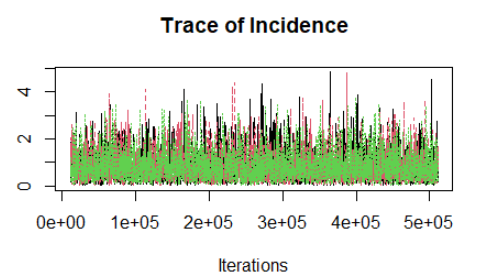

**FIGURE 8** **Second-line prophylaxis cohorts using total events**

**TABLE 2 Meta-analysis results for the AML cohorts**

| **Analysis subgroup** | **Number of studies** | **GLMM analysis** | | | **Beta-Binomial analysis** | |
| --- | --- | --- | --- | --- | --- | --- |
|  |  | **FE (95% CI)** | **RE (95% CI)** | **I^2^ (95% CI)** | **FE (95% CI)^1^** | **RE (95% CI)^2^** |
| Prophylaxis cohorts using total events | 3 | 1.16 (0.75, 1.79) | 1.16 (0.75, 1.79) | 0.00 (0.00, 89.6)  p=0.5925 | 1.17 (0.75, 1.83) | 1.23 (0.74, 1.86) |
| ^1^FE Beta binomial calculated using the built-in function in R (oad)  ^2^RE calculated via Bayesian approach using the WINBUGS code (pjpbayes.txt) via RJags. | | | | | | |

CI = confidence interval; FE = fixed effects; GLMM = generalized linear mixed model; RE = random effects

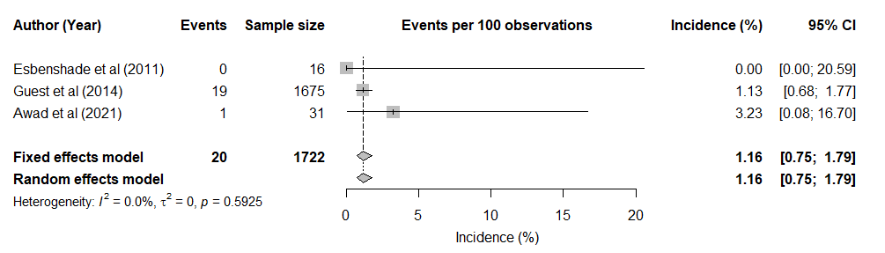

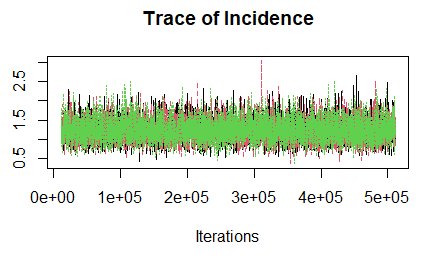

**FIGURE 9** **AML Prophylaxis cohorts using total events**

**TABLE 3 Meta-analysis results for other cancers cohorts**

| **Analysis subgroup** | **Number of studies** | **GLMM analysis** | | | **Beta-Binomial analysis** | |
| --- | --- | --- | --- | --- | --- | --- |
|  |  | **FE (95% CI)** | **RE (95% CI)** | **I^2^ (95% CI)** | **FE (95% CI)^1^** | **RE (95% CI)^2^** |
| No prophylaxis and confirmed events in the Neuroblastoma group | 4 | 3.33 (1.60, 6.83) | 3.33 (1.60, 6.83) | 0.00 (0.00, 84.7)  p=0.981 | 3.45 (1.62, 7.33) | 3.77 (1.68, 6.75) |
| No prophylaxis and total events in the Rhabdomyosarcoma group | 3 | 2.03 (0.91, 4.44) | 1.74 (0.47, 6.24) | 0.00 (0.00, 89.6)  p=0.9432 | 2.13 (0.58, 7.82) | 2.37 (0.96, 4.43) |
| No prophylaxis and total events in the Wilm’s tumour group | 3 | 0.81 (0.30, 2.14) | 0.76 (0.22, 2.62) | 1.1 (0.0, 89.7)  p=0.36 | 0.83 (0.28, 2.45) | 1.01 (0.32, 2.06) |
| ^1^FE Beta binomial is calculated using the built-in function in R (oad)  ^2^RE is calculated via Bayesian approach using the WINBUGS code (pjpbayes.txt) via RJags. | | | | | | |

CI = confidence interval; FE = fixed effects; GLMM = generalized linear mixed model; RE = random effects

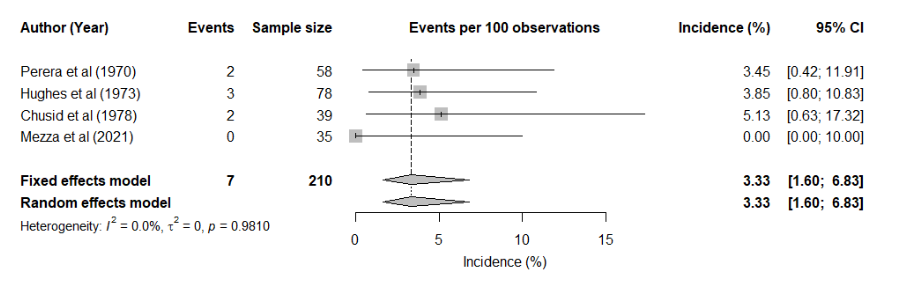

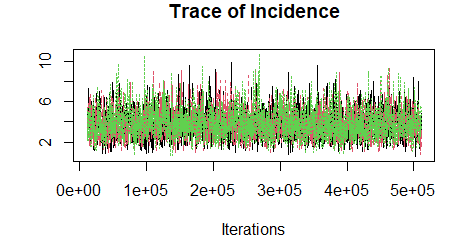

**FIGURE 10** **No prophylaxis using confirmed events - Neuroblastoma cohorts**

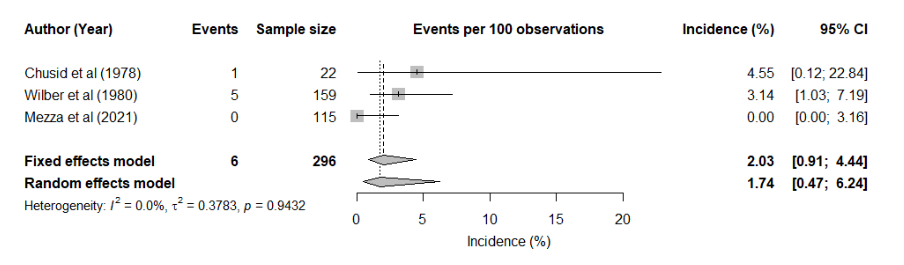

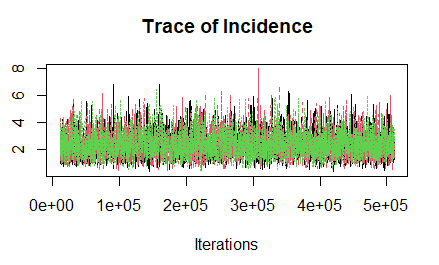

**Figure 11** **No prophylaxis using total events - Rhabdomyosarcoma cohorts**

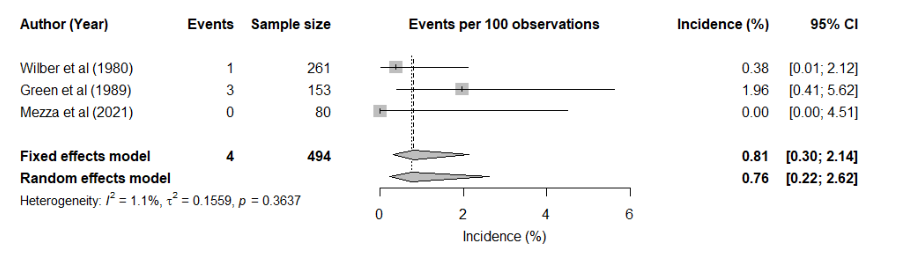

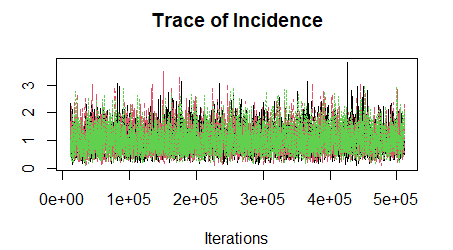

**FIGURE 12** **No prophylaxis using total events - Wilm’s tumour cohorts**
