## Supporting Results S3 for "A systematic review and meta-analysis of the incidence of *Pneumocystis jirovecii* pneumonia (PJP) in children and young people with cancer, cancer-like conditions or haematopoietic stem cell transplants"

### **Supporting Results S3. Summary of basic data extraction studies**

Prophylaxis studies of HSCT or ALL cohorts of less than 100 patients, or mixed population studies where PJP results were not reported by subpopulation were extracted using a basic data table (see Table 1 below).

### **Incidence of PJP**

#### **HSCT studies**

Fifteen HSCT studies were included in the basic data extraction, ranging in publication date from 1997 to 2020. Seven studies (47%) reported no PJP events among 256 CYP receiving a HSCT. Eight studies (53%) reported a total of 11 events among 347 CYP, with individual study incidence ranging from 1% (one event in 96 CYP) to 16.7% (two events in 12 CYP). Across the 15 studies, the total incidence of PJP was 1.8%.

#### **ALL-only studies**

Ten ALL-only studies were included, ranging in publication date from 1978 to 2020. Two studies (20%) reported no PJP events among 154 CYP. Eight studies (80%) reported a total of 17 events among 397 CYP, with individual incidence ranging from 1.7% (one event in 60 CYP) to 16.7% (six events in 36 CYP). Across the ten studies, the total incidence of PJP was 3.1%.

#### **Mixed cancer studies**

Eighteen mixed cancer studies, ranging in publication date from 1977 to 2021 were included in the basic data extraction. Six studies (33%) reported no PJP events among 584 CYP. Ten studies reported (56%) a total of 41 events among 14,131 CYP, with individual study incidence ranging from 0.01% (one event in 9675 CYP) to 10.6% (17 events in 1160 CYP). For two studies, either the number of patients with PJP or total number of patients assessed was not reported so incidence proportion could not be calculated. Across the 16 studies reporting both a numerator and denominator, the total incidence of PJP was 0.3%.

**TABLE 1 Studies with basic data extraction - HSCT and ALL prophylaxis studies of n<100; mixed population studies with PJP results not reported by subpopulation**

| **Author (Year), country** | **Number of patients** | **Study design** | **Population** | **Prophylaxis type (% patients)** | **Rate of PJP infection** |
| --- | --- | --- | --- | --- | --- |
| **ALL-only studies N<100** | | | | | |
| Agrawal (2011), USA^1^ | 87 | Retrospective cohort | 100% ALL; mean age 7 years; 51% male | TMP/SMX (100%) | 0/87 (0%) |
| Arico (1992), Italy^2^ | 67 | RCT | 100% ALL; 60% male | Co-trimoxazole daily (52%), co-trimoxazole 3 days consecutively per week (48%) | 0/67 (0%) |
| Goorin (1985), USA^3^ | 60 | RCT | 100% ALL; median age 5 years; 63% male | TMP/SMX (50%), placebo (50%) | 1/60 (1.7%). The single case was in the placebo group who subsequently died due to PJP. |
| Harris (1998), USA^4^ | 52 | Single-arm clinical trial | 100% ALL (in first relapse) | NR | 1/52 (1.9%) |
| Hewson (2020), UK*^5^ | 12 | Retrospective cohort | 100% ALL | 2^nd^ line after co-trimoxazole. At least 50% had Dapsone | 1/12 (8.3%) |
| Nakamura (2000), USA^6^ | 72 | Retrospective cohort | 100% ALL; average age 6 years | TMP/SMX (NR) | 2/72 (2.8%) confirmed - 1 patient died from PJP. Further 6 suspected cases of PJP. |
| Wolff (1978), USA^7^ | 36 | RCT | 100% ALL; 47% male | TMP/SMX (53%), no prophylaxis (47%) | 6/36 (16.7%). 2/19 (11%) for prophylaxis vs 4/17 (24%) for no prophylaxis |
| Camitta (1980), USA^8^ | 54 | RCT | 100% ALL; age range 1-15 years; 52% male | “Bactrim” i.e. TMP/SMX (only prescribed after 3 cases of PJP were identified) | 3/54 (5.6%). All cases were in the intensive phase group (3/28, 10.7% vs 0/26, 0% for non-intensive phase group) – 1 of these patients died from PJP infection. |
| Nazir (2017), Egypt^9^ | 62 | “Retrospective study with prospective follow-up” | 100% ALL; at least 52% male | TMP/SMX (39%), dapsone (55%), pentamidine (5%), atovaquone (1%) | 2/62 (3.2%) – one received TMP/SMX (1/24, 4.2%); one received dapsone (1/34, 2.9%) |
| Tsai (1985), Taiwan (not reported in English)^10^ | 49 | NR | 100% ALL | NR (received TMP/SMX as treatment) | 1/49 (2%) |
| **Mixed cancer studies** | | | | | |
| Weinthal (1994), USA^11^ | 22 | Retrospective cohort | 82% ALL, 18% ANLL; mean age 6.9 years; 68% male | Second-line aerosolised pentamidine (100%) (intolerant to TMP/SMX) | 0/22 (0%) |
| Rossi (1987), Italy^12^ | 90 | RCT | 63% ALL, 19% AML, 18% leukaemic relapse (NOS); Median ages 8 years and 5.5 years | TMP/SMX continuous (49%) vs TMP/SMX 3-days a week (51%) | 2/90 (2.2%) - 1/44 (2.3%) for TMP/SMX continuous vs 1/46 (2.2%) for TMP/SMX 3-days a week |
| Madden (2007), USA^13^ | 86 | Retrospective cohort | 66% ALL, 31% AML, 1% JCML, 1% refractory anaemia; mean age 10.7 years; 56% male. 43% received allogeneic HSCT | Second-line dapsone (100%) (intolerant to TMP-SMX) | 0/86 (0%) |
| Sartori (1993), UK^14^ | 12 | NR | 92% ALL, 8% NHL; median age 5 years; 50% male | Nebulised pentamidine (100%) | 1/12 (8.3%) |
| Zwaan (2017), International^15^ | 28 | Single-arm, phase 4 clinical trial | 61% ALL r/r, 39% LBL; mean age 11.5 years; 71% male | NR but assumed based on study year | 1/28 (3.6%) |
| O’Sullivan (1994), USA^16^ | 9 | Case series | 89% ALL, 11% AML; mean age 7.3 years. Unable to tolerate TMP/SMX | Pentamidine (100%) | 0/9 (0%) |
| Christensen (2005), Denmark^17^ | 98 (N=66 had fever) | Prospective cohort | NR for 98. For 66 with fever: 61% haematological cancers, 39% solid tumours; 55% male; 100% patients admitted with fever | NR | 1/98 (1.5%) |
| Wennmann (2021), Germany^18^ | 8 of interest | Retrospective cohort | 75% oncological disease, 25% HSCT. 100% received rituximab | Co-trimoxazole (% NR) | 0/8 (0%) |
| Proudfoot (2021), UK^19^ | NR | Prospective survey | NR | NR | 32 cases in 31 patients - 19 cases proven (8 ALL, 5 AML, 3 NHL, 2 neuroblastoma, 1 Ewings sarcoma) |
| Hughes (1977), USA^20^ | 160 | RCT | 85% ALL; 66% male | TMP/SMX (50%), placebo (50%) | 17/160 (10.6%) – all cases were in the placebo group |
| Kavcic (2013), USA^21^ | 434 | Retrospective cohort | Mixed malignancies; Median age 11 years; 71% male | NR but assumed based on study year | 0/434 (0%) |
| Orgel (2014), USA^22^ | 312 | Retrospective cohort | 50% ALL, 27% brain tumours, 23% other malignancies; Mean age 9.3 years; 63% male | Pentamidine (100%) | 3/312 (1%) confirmed (3 patients with ALL). Further 2 suspected cases (1 neuroblastoma, 1 high grade glioma) |
| Prasad (2008), USA^23^ | 223 | Retrospective cohort | 70% ALL, 3% AML, 8% Hodgkins lymphoma, 11% NHL, 5% solid tumours, 3% HSCT; Median age 10 years; 67% male | TMP/SMX (64%), Dapsone (16%), Pentamidine (15%), Atovaquone (2%), none (3%) | 4/223 (1.8%) – all 4 received prophylaxis, 2 ALL, 1 AML, 1 rhabdomyosarcoma |
| Goetz (1981), Germany^24^ | 200 | Prospective cohort | 45% leukaemia, 55% solid tumours; Mean age 9 years | NR but assumed not based on study year | 7 events of PJP (not clear how many patients) |
| Hughes (1984), USA*^25^ | 9,675 | Retrospective cohort | Paediatric oncology patients 100% | TMP/SMX (100%) | 1/9675 (0.01%). Data extracted digitally from graph |
| Shaw (1992), UK^26^ | 219 | Retrospective cohort | 65% leukaemia, 35% solid tumours | TMP/SMX (% NR) | 2/219 (0.9%) – number of episodes reported |
| Savasan (2021), USA^27^ | 25 | Prospective cohort | 28% HSCT, 72% chemotherapy-patients; median age 8 years; 80% male | Intravenous pentamidine (100%) | 0/25 (0%) |
| Wilber (1980), USA^28^ | 3314 (reported between 1977-1979) | Retrospective cohort | Paediatric oncology patients 100% (1976-1979 cohort) | TMP/SMX (% NR) | 9/3314 (0.27%) – 8 with ALL, 1 NHL 2 died |
| **HSCT studies N<100** | | | | | |
| Chaudhury (2020, USA*^29,30^ | 15 | Single-arm clinical trial | Allogeneic HSCT. 100% cGVHD. Median age 11 years | NR but assumed based on study year | 1/15 (7%) |
| Kruizinga (2017), Netherlands^31^ | 96 | Retrospective cohort | 96% allogeneic, 4% autologous | Pentamidine (100%) -35% first line, 65% second line | 1/96 (1%) – patient received second-line pentamidine |
| Maltezou (1997), USA^32^ | 32 (33 HSCTs) | Retrospective cohort | 76% allogeneic, 24% autologous; median age 10 years | Dapsone (100%) | 0/32 (0%) |
| Montoya Tomayo (2017), Spain^33^ | 55 (92 HSCTs) | Retrospective cohort | 100% autologous; median age 4.3 years; 65% male | Pentamidine (100%) | 0/55 (0%) |
| Nazir (2020), Oman^34^ | 12 | Retrospective cohort | 100% allogeneic; mean age 11.7 years; 42% male | Co-trimoxazole (100%) | 2/12 (16.7%) – both cases interrupted prophylaxis due to intolerance |
| Gonzalez Vicent (2019), Spain^35^ | 22 | Single-arm clinical trial | 100% allogeneic; 100% steroid-refractory-GVHD; median age 11 years; 45% male | Co-trimoxazole (100%) | 1/22 (4.5%) |
| Grunebaum (2006), Canada and Italy^36^ | 94 | Retrospective cohort | 100% allogeneic | NR but assumed based on study year | 3/94 (3.2%). 2 patients died from PJP |
| Neven (2019), France^37^ | 27 | Prospective cohort | 100% allogeneic; median age 1.5 years; 67% male | TMP/SMX (100%) | 1/27 (3.7%) – developed PJP 9 months post-HSCT whilst off prophylaxis |
| Nishimura (2021), Japan^38^ | 42 | Retrospective cohort | 100% allogeneic; median age 2 years; 83% male | NR but assumed based on study year | 0/42 (0%). 7 patients had experienced PJP pre-transplant |
| Pluchart (2012), France^39^ | 28 (35 HSCTs) | Retrospective cohort | 100% allogeneic; median age 7 years; 61% male | NR but assumed based on study year | 1/28 (3.6%) – developed PJP infection 9 months post-HSCT |
| Scott (2017), Canada^40^ | 14 | Retrospective cohort | 100% allogeneic; median age 5.4 months; 57% male | NR but assumed based on study year | 0/14 (0%) |
| Yang (2021), China^41^ | 53 | Retrospective cohort | 100% allogeneic; median age 7years; 64% male | NR but assumed based on study year | 1/53 (1.9%) – this patient had cGVHD |
| Brown (2013), USA^42^ | 43 | Retrospective cohort | 100% autologous; median age 4.1 years; 53% male | Either TMP/SMX, dapsone, atovaquone or pentamidine (% for each NR) | 0/43 (0%) |
| Nageswara Rao (2012), USA^43,44^ | 12 (13 HSCTs) | Retrospective cohort | 100% allogeneic; median age 8.1 years; 67% male | As per institutional standards (% NR) | 0/12 (0%) |
| Machatschek (2003), USA^45^ | 58 | Retrospective cohort | 100% autologous; median age 7.9 years; 50% neuroblastoma | TMP/SMX (% NR) | 0/58 (0%) |

* A study published as a conference abstract/letter to editor

ALL = acute lymphoblastic leukaemia; AML = acute myelocytic leukaemia; ANLL = acute non-lymphoblastic leukaemia; BAL = bronchoalveolar lavage; cGVHD = chronic graft-versus-host disease; EBV-PTLD = EBV post-transplant lymphoproliferative disorder; HSCT = haematopoietic stem cell transplant; LBL = lymphoblastic lymphoma; MUD = matched unrelated donor; MMRD = mismatched related donor; NHL = non-Hodgkin’s lymphoma; NOS = not otherwise specified; NR = not reported; PJP = pneumocystis jirovecii pneumonia; RCT = randomised control trial; RID = related HLA-identical donor; r/r = relapsed/refractory; SCID = severe combined immune deficiency; SR-GVHD = steroid refractory graft-versus-host disease; TMP/SMX = trimethoprim/sulfamethoxazole prophylaxis; USA = United States of America
