## Supporting Results S4 for "A systematic review and meta-analysis of the incidence of *Pneumocystis jirovecii* pneumonia (PJP) in children and young people with cancer, cancer-like conditions or haematopoietic stem cell transplants"

### Results for other outcomes

Deaths due to PJP were not widely reported, with small numbers of cases in each series where mortality was explicitly stated. Rates ranged from 0 to 100% and study reporting varied, with some reporting deaths based on confirmed PJP events and others reporting deaths based on the total PJP events (i.e. studies where there was some uncertainty about the validity of PJP diagnosis).

Only three HSCT cohorts reported timing of PJP diagnosis, each for one patient with PJP. Timings ranged from 192-198 days after 1^st^ HSCT in two cases and at 15 days after 2^nd^ HSCT in another CYP. The timing of PJP was reported in 13 ALL cohorts. In most cases, PJP occurred soon after diagnosis or after starting treatment (be it induction, consolidation or maintenance) and it was less common to have occurred after completing treatment/during remission. Similarly, across the mixed cancer studies, timing of PJP was poorly reported (only five cohorts) but again occurred early during the treatment period.
